# Hidden Cities, Hidden Gaps: Measuring Facility Readiness for Maternal and Newborn Health Services and its Association with Person-Centered Maternity Care in Urban Informal Settlements of Nairobi, Lusaka and Ouagadougou cities

**DOI:** 10.1101/2025.03.04.25323334

**Authors:** Safia S. Jiwani, Martin Mutua, Kadari Cisse, Choolwe Jacobs, Anne Njeri, Mwiche Musukuma, Godfrey Adero, Ashley Sheffel, Melinda K Munos, Elizabeth Stierman, Cheikh Faye, Ties Boerma, Agbessi Amouzou

## Abstract

**Background:** In sub-Saharan Africa, maternal and newborn deaths remain disproportionately higher among low-income populations, and they are associated with delivery in poorly equipped facilities and a shortage of staff to manage birth complications. We measured facility readiness to provide essential maternal and newborn health services and its association with women’s experience of person-centered maternity care (PCMC), and we compared facilities serving and not serving informal settlements in Nairobi, Lusaka and Ouagadougou cities.

**Methods:** We conducted a health facility assessment in public and private facilities serving select urban informal settlements in Nairobi, and we used existing data in Lusaka and Ouagadougou. We computed readiness indices for labor and delivery care, and small and/or sick newborn care (SSNC) in each city, and used t-tests to compare them across facilities serving and not serving informal settlements. We linked women’s self-reported PCMC scores to the labor and delivery readiness score of the facility they attended and ran 2-level linear regression models testing the association between facility readiness and PCMC scores.

**Results:** Facility readiness scores were computed among 18, 38 and 138 facilities offering delivery services in Nairobi, Lusaka and Ouagadougou respectively. Mean labor and delivery readiness scores in facilities serving informal settlements ranged from 55.9% in Ouagadougou to 73.6% in Lusaka; SSNC readiness ranged from 37.2% in Ouagadougou to 61.3% in Nairobi.

While facilities serving informal settlements had statistically significantly poorer readiness in Lusaka and Ouagadougou, key items such as newborn caps, registers, guidelines, and staff trained in Kangaroo Mother Care were lacking across both areas. We found no significant association between facility readiness and PCMC.

**Conclusions:** All facilities have substandard readiness for essential maternal and newborn health services, but those serving informal settlements are more disadvantaged. Investments in service readiness and quality of care remain critical.

## Background

The rapid growth of cities has been identified as one of the most pressing global health issues of the 21^st^ century.(1) In sub-Saharan Africa (SSA), 6 in 10 people are projected to live in cities by 2050.(2) Large rural-to-urban migration has led to the expansion of urban informal settlements, threatening governments’ capacity to ensure universal access to essential health services. While urban living exposes individuals to better employment, education, health and living conditions, the urban advantage is not equitably distributed among urban residents. This has created “hidden cities”(1) of urban poor populations that no longer benefit from the urban advantage.(3,4)

The challenges posed by rapid urbanization are reflected in maternal and newborn health outcomes:(5,6) a staggering 70% of global maternal deaths and almost a third of livebirths occur in sub-Saharan Africa,(7,8) and evidence suggests that most deaths in LMICs are due to poor quality of care, rather than inadequate access to services.(9) Most maternal and newborn deaths occur during the intrapartum and immediate postpartum period, and are associated with unskilled birth attendance, delivery in poorly equipped facilities and a shortage of essential obstetric care supplies to manage birth complications.(10) Birth asphyxia, preterm births and infections account for 87% of newborn deaths in SSA,(11) and can be prevented by ensuring access to high quality life-saving interventions,(12) such as thermal protection of preterm babies and specialized treatment for newborns born small (premature or with low birthweight) or sick.(13) However, these services are not equitably available to all populations in need, and their quality remains variable.

Measuring quality of care, a complex multidimensional concept that goes beyond mere access to services, is therefore essential to identify gaps and target interventions effectively. The World Health Organization’s quality of care framework for maternal and newborn health,(14) stemming from the Donabedian model,(15) encompasses three core dimensions: provision of care, experience of care, and structural quality, also known as facility readiness. These dimensions not only shape health outcomes but also influence the long-term trust and engagement of urban populations with the healthcare system.

Facility readiness is defined as the capacity of a health facility to provide services following standard guidelines, based on the observed availability and functionality of key items such as equipment, commodities, amenities, standard precautions, the presence of trained staff, as well as appropriate systems in place to support high quality of care and safety.(14) It reflects cross-cutting physical and human resources and is a crucial pre-requisite for enabling high quality services. Facility readiness is generally measured through a facility inventory in health facility assessments (HFA), such as the DHS program’s Service Provision Assessment (SPA)(16) focusing on antenatal, family planning, maternity care and sick child services, as well as the WHO’s Service Availability Readiness Assessment (SARA)(17) and Harmonized Health Facility Assessment (HHFA)(18) covering a wider range of health services.

Patient experience of care is another core quality of care dimension and is typically measured through client exit surveys conducted upon discharge from facility care. Person-centered maternity care (PCMC) is one such measure referring to maternity care that is respectful of and responsive to women and their families’ preferences, needs and values, and ensuring that their values guide clinical decisions.(19) Afulani et al. have developed and validated a 30-item PCMC scale covering elements of patient dignity and respect, communication and autonomy, and supportive care.(20,21)

While quality of care is generally explored nationally or sub-nationally, little is known about the quality of services among the most vulnerable urban populations, and differences within urban areas. Our study objectives were to 1) measure facility readiness for labor and delivery care and small and/or sick newborn care (SSNC) services in Nairobi, Lusaka and Ouagadougou’s urban informal settlements, 2) compare service readiness between facilities serving and not serving urban informal settlements in Lusaka and Ouagadougou, and 3) assess the associations between labor and delivery care readiness and PCMC in informal settlements in each city. We hypothesized that women attending a facility with higher service readiness report a higher PCMC experience.

## Methods

### Study setting

This study was conducted within select informal settlements in Nairobi, Lusaka and Ouagadougou cities. These were locally defined geographic areas housing the urban poorest women in each city. Informal settlements dwellers typically lack tenure security, basic services and infrastructure, and their housing does not comply with safety regulations, leading to hazardous living conditions.(22) In Nairobi, we selected the Korogocho and Viwandani urban informal settlements based on existing research activities;(23) in Lusaka and Ouagadougou, the existing health facility data covered all facilities including those serving and not serving the cities’ informal settlements. Importantly, each context was unique and the characteristics of informal settlements and their residents in Nairobi differed from those in Lusaka or Ouagadougou. Details on the study setting in each city are reported elsewhere (REF: paper 1 under review).

### Study design and data

We conducted a study evaluating the quality of maternal and newborn health services in select urban informal settlements in Nairobi, Lusaka and Ouagadougou cities. We administered in-depth interviews with health providers, client exit surveys of women discharged from childbirth care, and health facility assessments for maternal and newborn health services between January and March 2023. This analysis focused on the health facility assessments, though we also used data from client exit surveys.

In Nairobi, we conducted a cross-sectional survey of health facilities offering maternal and newborn health services and primarily serving the Korogocho and Viwandani informal settlements. In Ouagadougou and Lusaka, we used existing data from the WHO’s Harmonized Health Facility Assessment (HHFA)(18) conducted in 2020/2021 and 2021/2022 respectively. Aside from minor country-specific adaptations, the variables were identical across countries. The HHFA are comprehensive health facility surveys that measure service availability and the capacity of facilities to meet standards of quality; they typically cover modules relating to service availability, service readiness, quality of care and management/finance. In Lusaka and Ouagadougou, these HHFA were conducted in a census of all health facilities, including those serving and not serving urban informal settlements.(24,25) In Lusaka, we defined facilities serving informal settlements as those located within an informal settlement; in Ouagadougou, given the small number of facilities located within informal settlements, they were defined as those located within a 1km radius of “*non-loti*” (informal) areas.

As part of this study, we also administered client exit surveys to a random sample of 1,249 women of reproductive age (15-49 years) discharged from childbirth care in health facilities serving a subset of urban informal settlements in Nairobi, Lusaka and Ouagadougou. The survey included a validated 30-item PCMC scale covering dignity and respect, communication and autonomy, and supportive care subscales.(20) Additional details on the design and sampling of client exit surveys are reported elsewhere (REF: paper 1 under review).

### Tool development and training

In Nairobi, we adapted the HHFA tool (version 1, September 2022) to the Kenyan context and restricted it to the following modules relevant for maternal and newborn health services: staffing and resources, infection prevention, outpatient maternal and newborn health services, delivery and newborn care services, laboratory, consumable commodities, pharmaceutical commodities. The tool was administered on a tablet and programmed using Survey CTO. Four clinicians were trained on the data collection process, and the tool was pre-tested during the pilot study. In Lusaka and Ouagadougou, the complete HHFA tool covering all service areas was implemented by WHO and in-country partner institutions.(24,25)

### Sample size

All functional health facilities of allopathic medicine (public, private for-profit and private non-profit/religious) offering maternal and newborn health services were eligible for the study. In Nairobi, we restricted the sample to facilities serving the Korogocho and Viwandani informal settlements, defined based on country classifications. A mapping exercise and formative research identified 43 eligible health facilities located within or immediately neighboring the informal settlements, of which 38 agreed to participate. From the HHFA data, we identified 115 facilities serving and 215 facilities not serving informal settlements in Ouagadougou, and 31 facilities serving and 88 facilities not serving informal settlements in Lusaka. We excluded tertiary-level referral and specialized hospitals, as there were very few and they served the whole city, to allow for a better comparison between facilities serving and not-serving informal settlements in Lusaka and Ouagadougou.

### Measures

#### Maternal and newborn health service readiness

This analysis focused on facility readiness for normal deliveries, as well as routine and special newborn care for small and/or sick newborns, reflecting the basic capacity needed in primary care facilities before referral to higher-level ones. Service readiness was defined based on the availability and functionality of key items needed to offer high quality care. We generated binary variables for each item, with a value of 1 if the item was observed available and functional on the day of the survey, and 0 otherwise. For labor and delivery care readiness, we generated a simple additive readiness score based on the number of items available and functional to provide high-quality service; each item therefore carried equal weight towards the overall score. Item selection was based on HHFA definitions, and additional items reflecting the WHO standards for improving maternal and newborn care,(26,27) informed by Stierman et al.(28) Our labor and delivery readiness index included 55 items covering the following five domains: equipment, medicines and commodities, basic amenities, human resources and guidelines, and performance of basic emergency obstetric and newborn care (BEmONC) signal functions in the last 12 months (supplementary table 1).

For SSNC readiness, we generated a weighted additive readiness score whereby each domain (rather than each item) was weighted equally towards the overall score. This was deemed more appropriate than a simple additive method given differing number of items per domain. We adapted the SSNC readiness index developed by Sheffel et al.,(29) based on their mapping and extraction of the 2020 WHO Standards for improving the quality of small and sick newborns care,(30) Survive and Thrive,(31) and works by Moxon et al.(32) The SSNC index included 50 items covering the following eight intervention domains, as well as a general facility readiness domain: immediate newborn care and routine care (including newborn drying, immunization, hygienic cord care, vitamin K, and eye care), early initiation and support for breastfeeding, neonatal resuscitation, prevention of mother-to-child transmission of HIV, Kangaroo Mother Care (KMC), detection and management of neonatal infection, comfort and pain management, and detection and management of hypoglycemia (supplementary table 2). Facilities that did not offer one or more of the above interventions received a value of 0 for all the corresponding domain items.

#### Person-centered maternity care

We estimated PCMC scores following Afulani et al.’s methodology, whereby a numeric value ranging from 0 to 3 is assigned to each of the 30 items making up the PCMC scale (20,21) (supplementary table 3). Responses are on a 4-point scale (no never, yes a few times, yes most of the time, yes all the time) and the highest value of 3 is attributed to the response option reflecting optimal person-centered behavior. Values for all 30 items are summed for a maximum score of 90 points for the full PCMC scale; a higher score therefore reflects higher PCMC. We also generated domain-specific scores for each of the three PCMC subscales: dignity and respect (6 items for a total of 18 points), communication and autonomy (9 items for a total of 27 points), and supportive care (15 items for a total of 45 points).(21)

### Statistical analysis

#### Maternal and newborn health service readiness

The analysis of service readiness was restricted to facilities offering labor and delivery care. We computed overall and domain-specific readiness scores for labor and delivery care as well as SSNC in each city. These were measured on a continuous scale ranging from 0 to 1 and were rescaled to a 100 for improved interpretability as a percentage. We evaluated floor and ceiling effects by reporting the proportion of facilities with very low (0-5%) or very high (95-100%) readiness scores: these were below 10% for both service areas, indicating adequate capacity to discriminate between facilities with very low/very high readiness. Additionally, we disaggregated readiness scores by facility type and managing authority. We used the same items to define service readiness in hospitals and lower-level facilities, reflecting the same standards for high quality care. However, given that some items such as infant incubators, resuscitation tables, x-ray machines and morphine were not expected to always be available in lower-level facilities, we also calculated the maximum score expected in these facilities when excluding such items.

In Lusaka and Ouagadougou, we compared mean readiness scores between facilities serving and not serving informal settlements using a t-test. We also compared the proportion of facilities with low (<60%), moderate (60-80%) and high (>80%) readiness scores; an arbitrary threshold of 80% was used to reflect facilities with most items available and functional.

#### Association between facility readiness and experience of person-centered maternity care

We conducted an exact match linking of individual women’s self-reported PCMC scores and the facilities’ labor and delivery care readiness scores, for each city separately. We linked the two separate data sources (the client exit survey and the health facility assessment) by facility, and we assigned to each woman’s self-reported PCMC score (from the client exit survey), the labor and delivery readiness score of the facility in which she gave birth (from the health facility assessment). The proportion of surveyed women linked to a facility was 97.3% in Lusaka, 99.3% in Nairobi and 100% in Ouagadougou. A subset of facilities were included in the linking analysis, given client exit surveys were conducted in a select informal settlements.

For each city, we ran a multilevel random intercept linear regression model of the PCMC score (out of 90) on the overall rescaled labor and delivery readiness score (%). In this multilevel model, level-1 referred to individual women and level-2 referred to the facility they attended, thus accounting for clustering of women within facilities and variability in PCMC scores across facilities. We ran crude and adjusted models, controlling for structural (women’s education and employment), intermediary (women’s age, marital status, parity, pregnancy complications, history of miscarriage/stillbirth, antenatal care) and health systems factors (facility type and managing authority, provider type assisting at birth, length of facility stay, receipt of maternal and newborn postnatal care and content of care prior to discharge). In the final model, we included the covariates that had bivariate associations with PCMC significant at p value < 0.2, and we assessed for multicollinearity using variance inflation factors. As supplementary analyses, we tested the association between labor and delivery readiness scores and each PCMC domain separately.

## Results

### Characteristics of facilities offering maternal and newborn health services

In Lusaka, 80.7% of facilities serving informal settlements were public health centers and 12.9% were public hospitals, whereas those not serving informal settlements were mostly private facilities. In Nairobi and Ouagadougou, 92% and 85.3% of facilities serving informal settlement respectively were health centers, the majority being private. Most facilities across cities had continuous power supply and access to an improved water source, however less than half of all facilities had an ambulance or a vehicle available 24 hours on the day of the survey (table 1).

**Table 1.** Characteristics of facilities offering maternal and newborn health services by city.

|  | LUSAKA |  | NAIROBI | OUAGADOUGOU |  |
| --- | --- | --- | --- | --- | --- |
|  | serving informal settlement (n=31) | not serving informal settlements (n=88) | serving informal settlements (n=38) | serving informal settlements (n=115) | not serving informal settlements (n=215) |
| **Facility type and managing authority\*** | (%) | (%) | (%) | (%) | (%) |
| Public health center/other | 80.7 | 30.7 | 21.0 | 27.0 | 26.5 |
| Private for-profit health center/ other | 6.4 | 45.5 | 50.0 | 40.9 | 32.6 |
| Private non-profit health center/ other | 0.0 | 13.6 | 21.0 | 17.4 | 7.4 |
| Public hospital | 12.9 | 4.5 | 7.9 | 1.7 | 6.5 |
| Private for-profit hospital | 0.0 | 5.7 | 0.0 | 6.9 | 19.5 |
| Private non-profit hospital | 0.0 | 0.0 | 0.0 | 6.1 | 7.4 |
| **Staffing** |  |  |  |  |  |
| Mean # nurses/midwives per health center (min-max range) | 25.4 (1-107) | 12.7 (1-65) | 8.7 (0-50) | 4.2 (0-24) | 2.9 (0-38) |
| Mean # nurses/midwives per hospital (min-max range) | 164.5 (90-287) | 75.2 (2-296) | 69.7 (28-140) | 14.1 (0-173) | 9.7 (0-177) |
| Mean # physicians/obstetricians per health center (min-max range) | 45.1 (0-190) | 19.6 (0-141) | 0.2 (0-2) | 2.1 (0-22) | 6.7 (0-52) |
| Mean # physicians/obstetricians per hospital (min-max range) | 310 (18-619) | 70.0 (1-236) | 38.7 (1-68) | 15.3 (0-59) | 21.7 (0-128) |
| **Number of beds** |  |  |  |  |  |
| Mean # beds (inpatient, observation and maternity, excluding delivery beds) per health center (min-max range) | 5.1 (0-40) | 7.7 (0-65) | 5.6 (0-30) | 8.7 (0-52) | 6.6 (0-29) |
| Mean # beds (inpatient, observation and maternity, excluding delivery beds) per hospital (min-max range) | 91.0 (17-205) | 18.0 (4-50) | 72.7 (0-200) | 14.4 (2-48) | 20.8 (0-207) |
| **Basic amenities** |  |  |  |  |  |
| Continuous power supply (last 7 days) | 74.2 | 77.3 | 65.8 | 66.1 | 65.1 |
| Access to improved water source | 87.1 | 90.9 | 84.2 | 85.2 | 84.7 |
| **Emergency transport** |  |  |  |  |  |
| Functional 24h ambulance or vehicle with driver | 32.3 | 48.9 | 31.6 | 21.7 | 26.5 |

| **Laboratory & Pharmacy capacity** |  |  |  |  |  |
| --- | --- | --- | --- | --- | --- |
| Availability of in-house laboratory for diagnostic testing (%) | 51.6 | 68.2 | 94.7 | 74.8 | 76.3 |
| Availability of in-house storage of pharmaceuticals (%) | 100.0 | 98.9 | 97.4 | 85.2 | 73.5 |

*\* Facility types are defined as follows: In Lusaka health center/other refers to urban health posts, health centers or clinics, and hospitals refer to first and second level hospitals. In Nairobi, health center/other includes facility levels 2 & 3, hospital includes health facility levels 4 & 5. In Ouagadougou health center/other includes « niveau 1 échelon 1 » facilities (cabinet médical, centre de santé et promotion sociale, cabinet de soins infirmiers, clinique d'accouchement, dispensaire isolée, infirmerie), hospital refers to « niveau 1 échelon 2 facilities » (Centre médical avec antenne chirurgicale, clinique, centre médical).*

Approximately one third (35.5%) of facilities offering MNH services in Lusaka’s informal settlements offered labor and delivery care; In Nairobi and Ouagadougou’s informal settlements, close to half did. The reported availability of basic emergency obstetric and newborn care (BEmONC) varied between 32.3% to 47.4% in informal settlements across cities, however less than 10% of facilities reported offering all seven BEmONC signal functions. Only 5.2% and 9.3% of facilities serving and not serving informal settlements respectively in Ouagadougou administered oxygen. KMC was available in less than 20% of facilities, with lower availability in informal settlements: in Ouagadougou’s informal settlements, only 8.7% of facilities offered KMC (supplementary table 4).

### Facility readiness for labor and delivery care

The overall mean readiness score for labor and delivery care in facilities serving informal settlements was 73.6% (SD=12.5) in Lusaka, 69.5% (SD=19.4) in Nairobi and 55.9% (SD=11.9) in Ouagadougou. It was statistically significantly higher in facilities not serving informal settlements in Lusaka and Ouagadougou at 85.6% (SD=9.2) and 60.3% (SD=12.7) respectively (figure 1, supplementary materials: figure 1 & table 5). Similarly, in both these cities, there was a higher proportion of facilities serving informal settlements with low (<60%) readiness scores, and a smaller proportion with high readiness scores (>80%). For instance, only 27.3% of facilities serving informal settlements in Lusaka had high readiness compared to 74.1% of facilities not serving informal settlements. In Ouagadougou, no facility serving informal settlements had a readiness score above 80% (figure 2).

**Figure 1.**
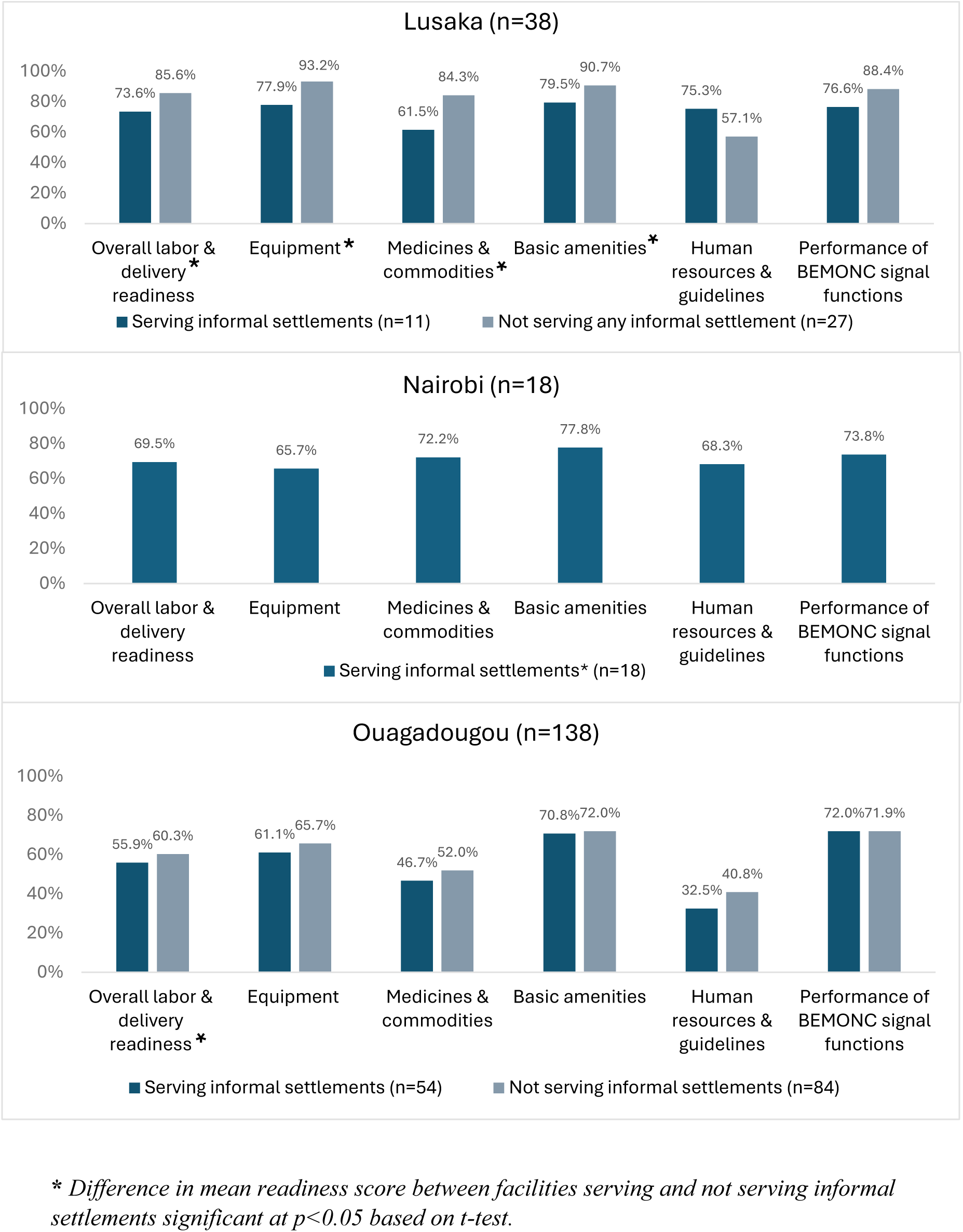
Labor & delivery care readiness scores (%) by domain and city.

**Figure 2.**
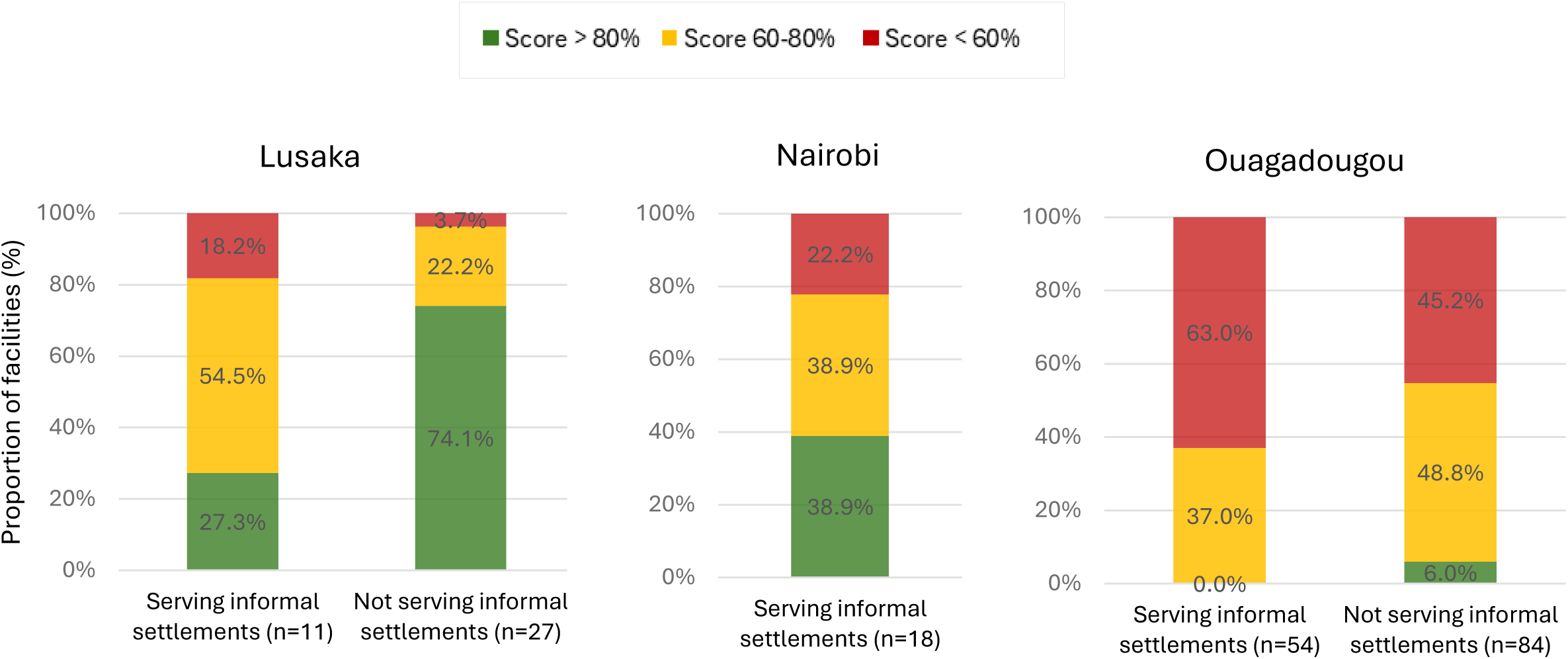
Proportion of facilities with high, moderate and low readiness scores for labor & delivery care.

Facilities serving informal settlements in Lusaka had statistically significantly poorer readiness scores for all domains except human resources and performance of BEmONC signal functions, and were least ready in terms of medicines and commodities with a domain score of 61.5% (figure 1, supplementary table 5). Key items that were lacking in Lusaka’s informal settlements included injectable vitamin K, hypertensive drugs, calcium gluconate and oxygen, in addition to basic equipment items (examination lights, vacuum aspirators and clean towels to dry newborns) (supplementary materials: figure 2A & table 6). In Ouagadougou, facilities serving and not serving informal settlements were deficient in the availability of trained human resources and guidelines, with domain scores of 32.5% (SD=25.0) and 40.8% (SD=26.4) respectively (figure 1, supplementary table 5). Checklists for essential childbirth care, staff trained in all areas of labor and delivery care, oxygen, misoprostol, and infant incubators were particularly unavailable in informal settlement facilities (supplementary materials: figure 2C & table 6). In Nairobi’s informal settlements, domain scores ranged from 65.7% (SD=23.8) for equipment to 77.8% (SD=19.0) for basic amenities (figure 1, supplementary table 5), though the following items generally had poor availability: vacuum aspirator, infant incubator, neonatal resuscitation bag and mask, and guidelines/checklists (supplementary materials: figure 2B & table 6).

When disaggregating labor and delivery readiness scores by facility type and managing authority, public hospitals serving informal settlements had higher readiness than their counterparts not serving informal settlements. In Lusaka, the four public hospitals in informal settlements had a higher overall readiness at 86.6% compared to 77.8% in the three public hospitals not serving informal settlements. In Ouagadougou, private facilities serving informal settlements, representing the majority of facilities serving these areas, were the least ready for labor and delivery care. Similarly, in Nairobi, private health centers received the lowest score of 62.6% and public hospitals received the highest score of 87.7% (figure 3, supplementary table 7). Across cities, none of the facilities serving informal settlements, except one public hospital in Lusaka, met the readiness threshold expected in lower-level facilities (supplementary figure 3).

**Figure 3.**
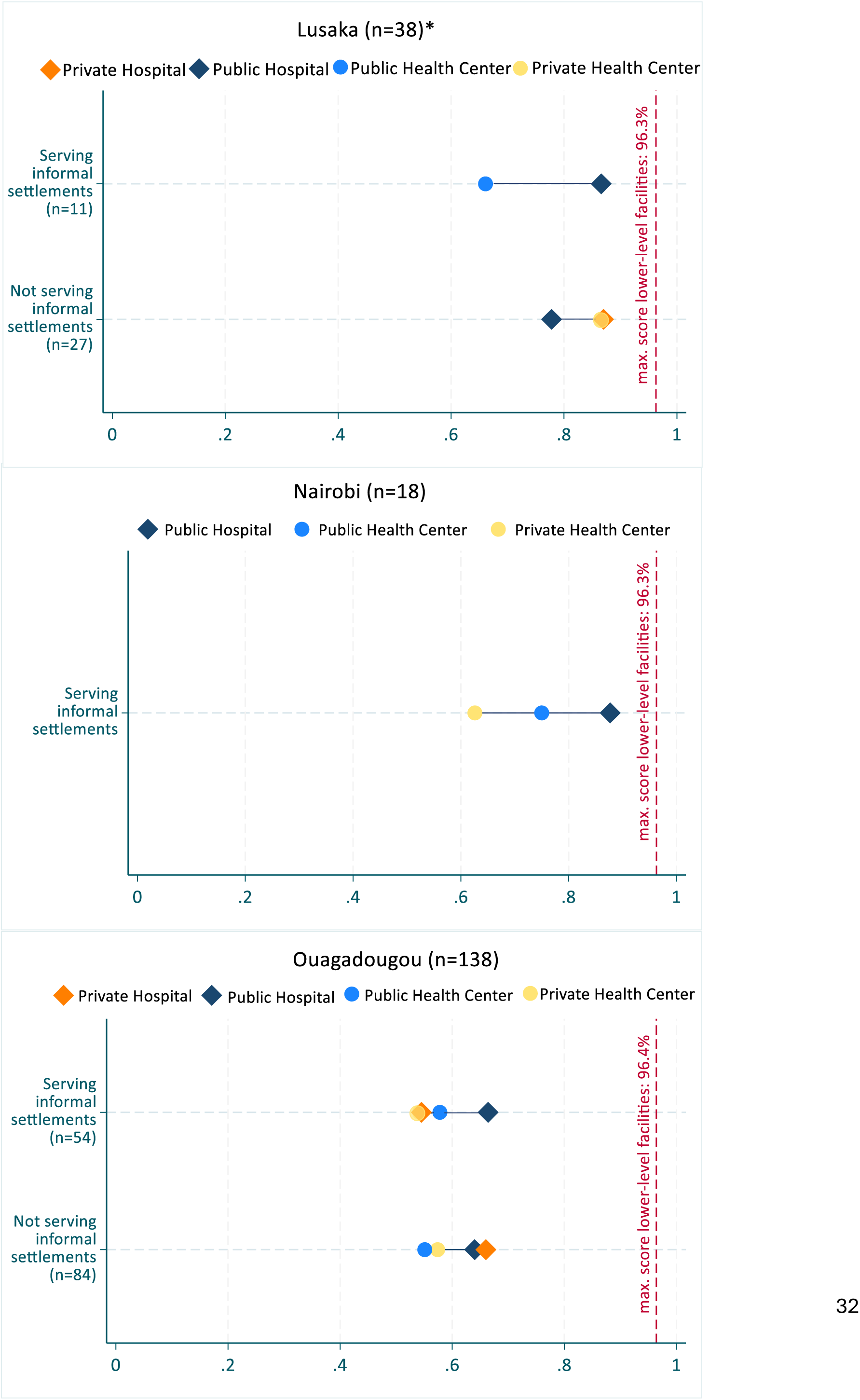
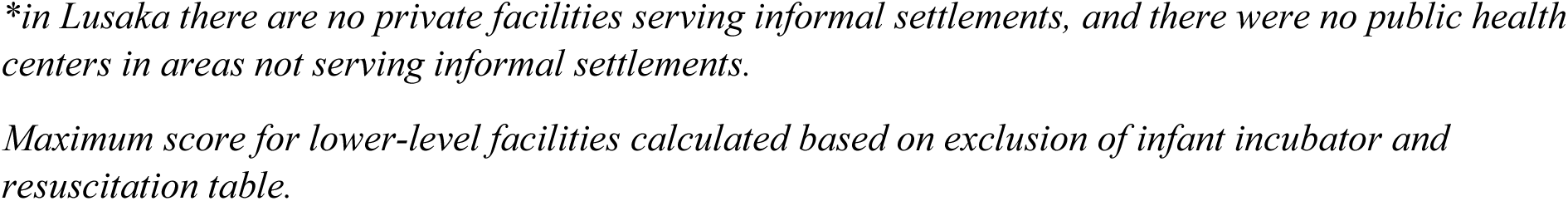
Labor & delivery care readiness scores (%) by facility type and managing authority.

### Facility readiness for small and sick newborn care

The overall mean SSNC readiness score in facilities serving informal settlements was 60.8% (SD=14.5) in Lusaka, 61.3% (SD=17.9) in Nairobi and much lower at 37.2% (SD=9.3) in Ouagadougou, and it was statistically significantly higher in facilities not serving informal settlements (70.7% in Lusaka and 40.3% in Ouagadougou) (figure 4, supplementary materials: table 5 & figure 4). In Ouagadougou, 98.1% and 92.9% of facilities serving and not serving informal settlements respectively had low readiness for SSNC (scores <60%), and no facility serving informal settlements scored over 80%. The latter was also observed in Lusaka, whereas 25.9% of facilities outside informal settlements scored over 80% (figure 5).

**Figure 4.**
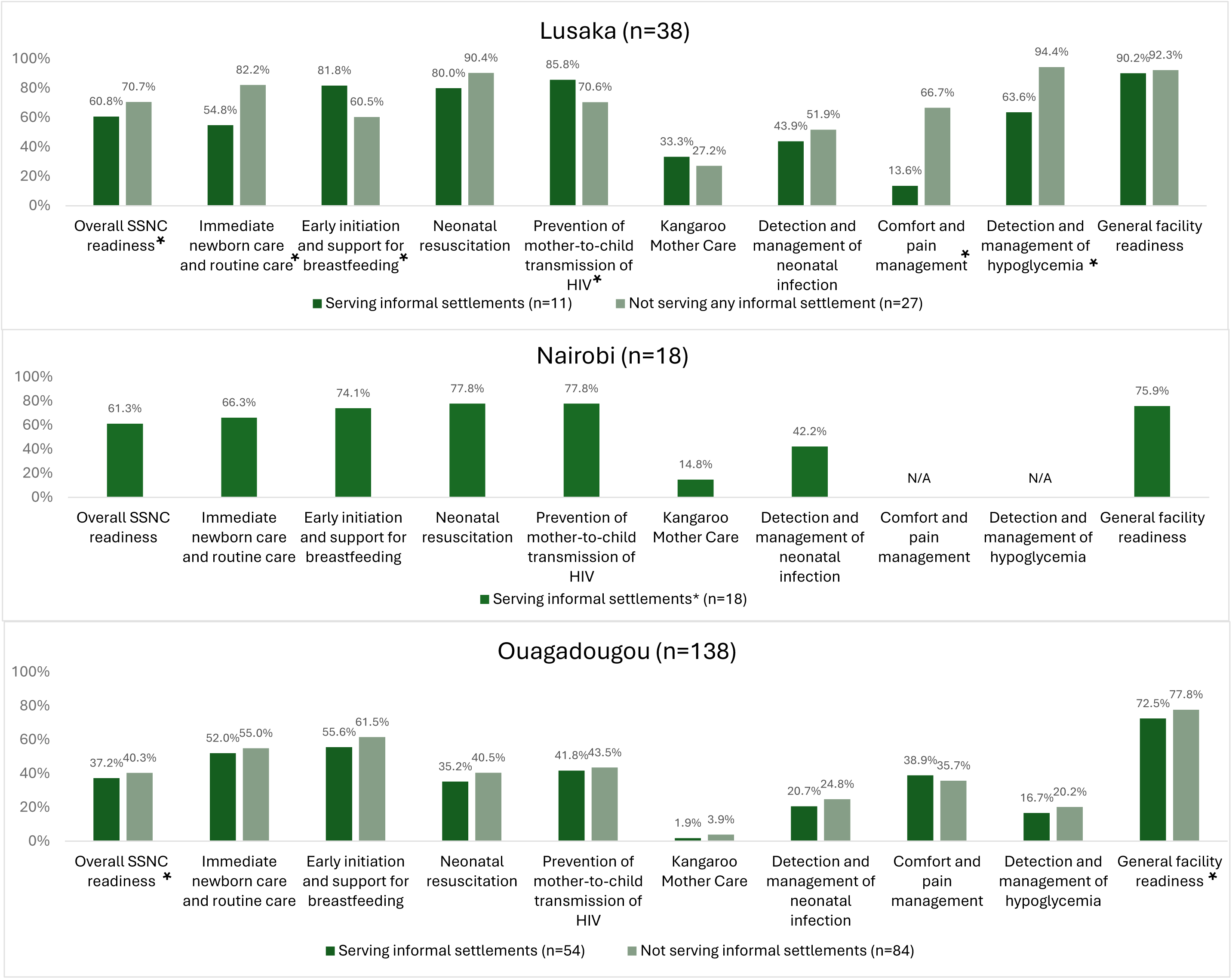
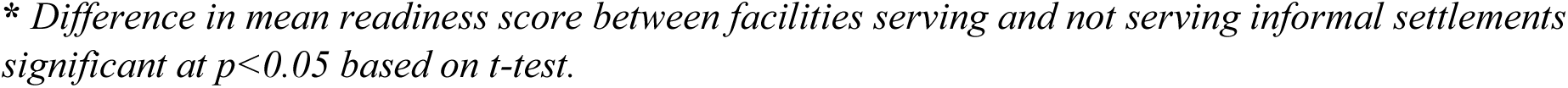
Small & sick newborn care readiness scores (%) by domain and city.

**Figure 5.**
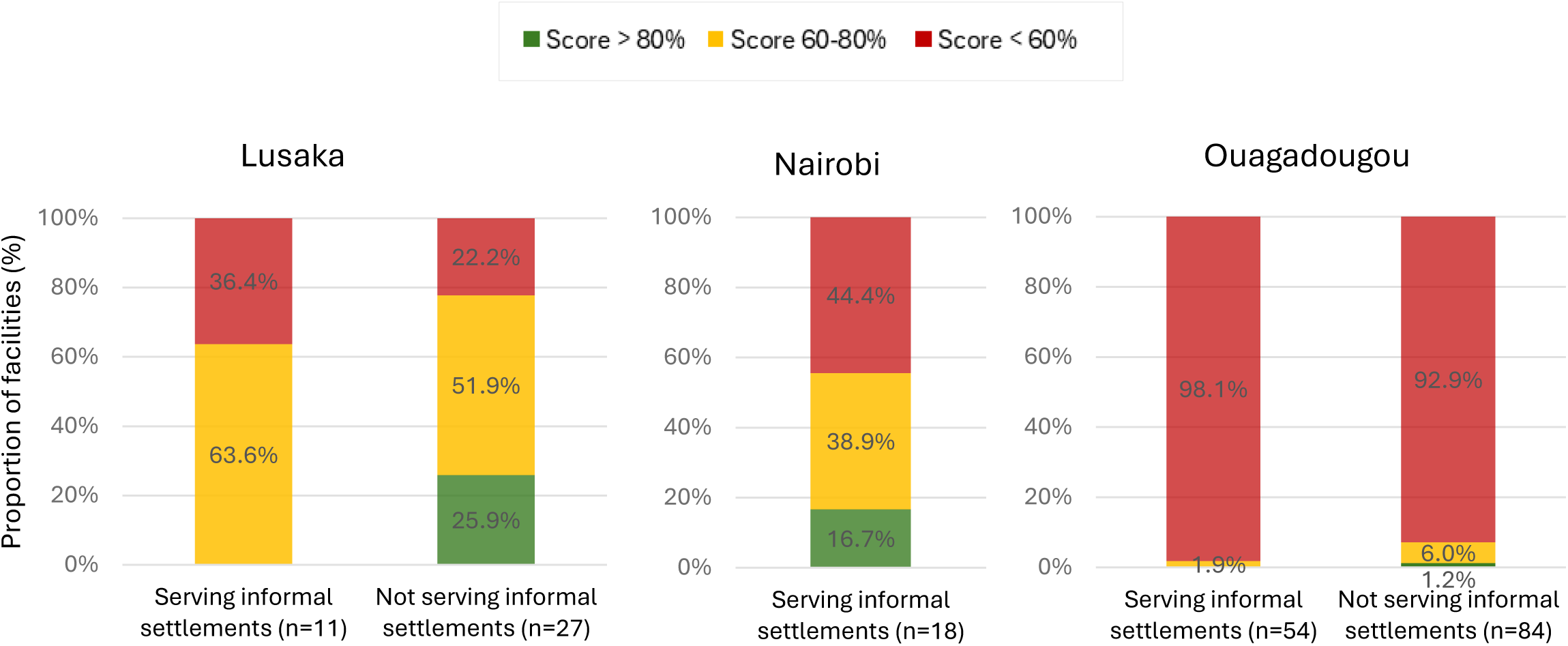
Proportion of facilities with high, moderate and low readiness scores for small and sick newborn care.

In Lusaka’s informal settlements, SSNC intervention domain scores ranged from 13.6% for comfort and pain management to 85.8% for PMTCT. KMC readiness was consistently poor across areas, with large variability across facilities. This was also found in Nairobi’s informal settlements where the KMC domain score was 14.8% (SD=29.1). In Ouagadougou, readiness for SSNC was generally poorer than in Lusaka and Nairobi. Intervention domain scores in informal settlements ranged from a meager 1.9% (SD=6.6) for KMC to 55.6% (SD=25.9) for early initiation of breastfeeding (figure 4, supplementary table 5). Further analyses pointed to the general unavailability of trained staff in KMC, as well as guidelines and registers for KMC and neonatal sepsis across all facilities, both serving and not serving informal settlements (supplementary materials: figure 5A-C & table 8).

Public hospitals serving informal settlements consistently had higher SSNC readiness than both public and private hospitals not serving informal settlements in Lusaka and Ouagadougou (figure 6, supplementary table 7). In all cities, none of the facilities serving informal settlements achieved the maximum readiness score expected in lower-level facilities (supplementary figure 6).

**Figure 6.**
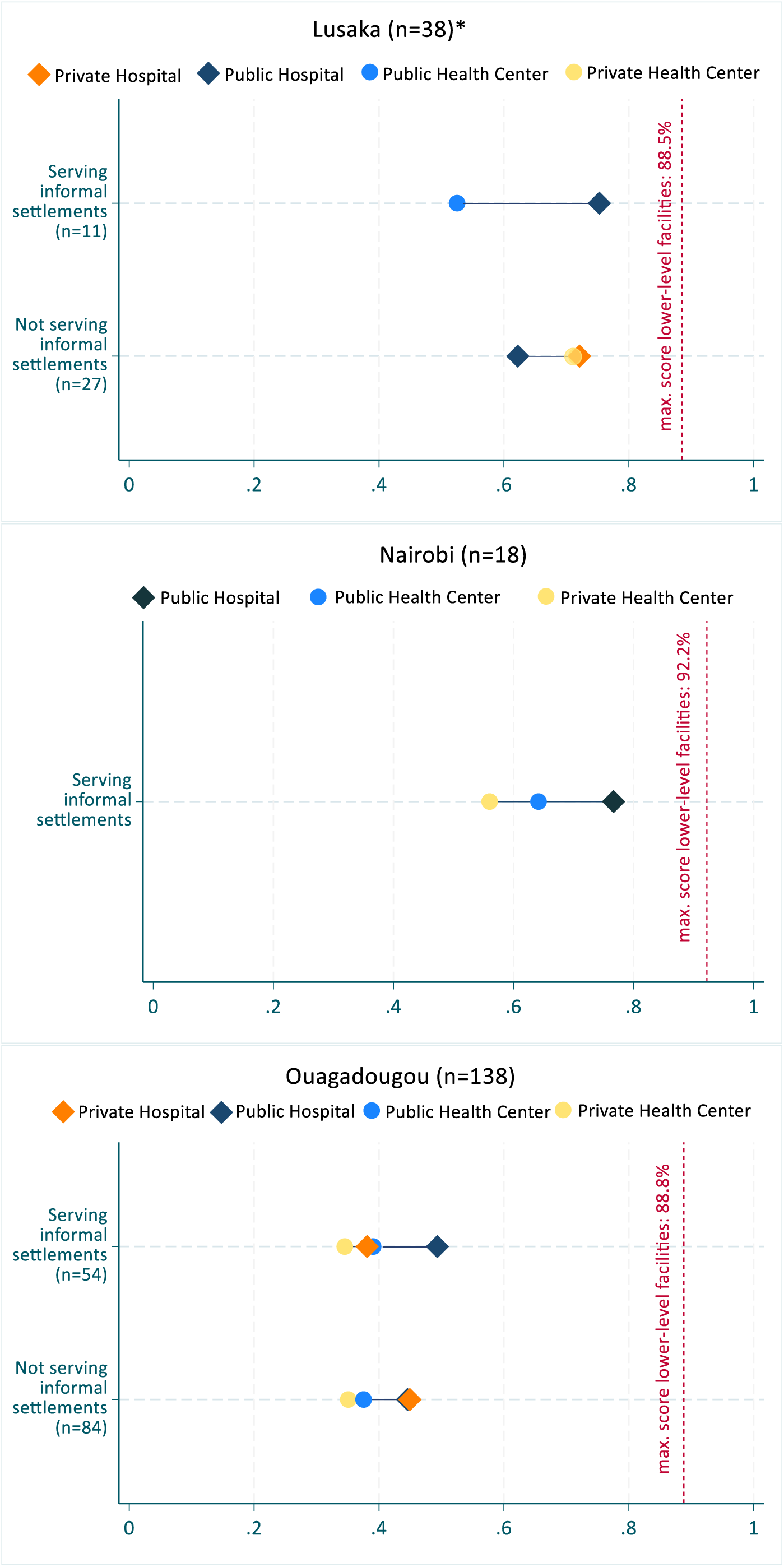
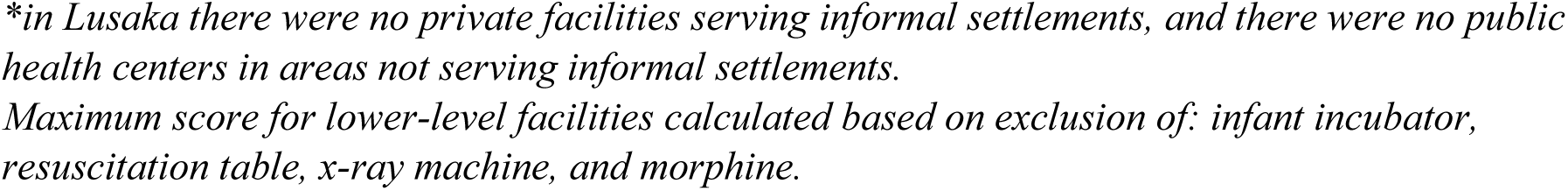
Small & sick newborn care readiness scores (%) by facility type and managing authority.

### Association between labor and delivery readiness and PCMC

In Lusaka, 424 women were linked to 10 facilities, in Nairobi 411 women were linked to 16 facilities, and in Ouagadougou 401 women were linked to 54 facilities. Most women were aged 20-34 years old, were unemployed, and were attended by a nurse or midwife during childbirth. In Lusaka and Nairobi, the majority of women gave birth in public hospitals, whereas in Ouagadougou they gave birth in public health centers (supplementary tables 9 & 10).

Multilevel models adjusting for relevant structural, intermediary and health systems determinants of PCMC suggested no statistically significant effect of giving birth in a facility with higher readiness on women’s overall PCMC experience (table 2). As a sensitivity analysis, we re-ran this model on a logit-transformed PCMC outcome, and the results were consistent. Our supplementary analyses for each PCMC domain separately indicated that, while statistically significant associations were observed, the magnitudes remained small: In Nairobi, a 10-point increase in facility readiness was associated with a 0.6-point decreased communication and autonomy score (out of 27 points), and a 0.8-point increased supportive care score (out of 45 points). In Lusaka, it was associated with a 2.2-point decreased supportive care score (supplementary table 11).

**Table 2.** Associations between facilities’ labor & delivery care readiness scores (%) and women’s PCMC scores (out of 90 points) in study areas by city.

|  | **Crude model** |  |  | **Adjusted Model** |  |  |
| --- | --- | --- | --- | --- | --- | --- |
|  | Coefficient<br>[95%CI] | P value | n | Coefficient<br>[95%CI] | P value | n |
| Lusaka<sup>+</sup> | -0.14<br>[-0.42,0.14] | 0.317 | 424 | -0.12<br>[-0.36,0.11] | 0.301 | 411 |
| Nairobi<sup>++</sup> | -0.04<br>[-0.25,0.18] | 0.725 | 411 | 0.04<br>[-0.10,0.17] | 0.579 | 406 |
| Ouagadougou<sup>+++</sup> | 0.05<br>[-0.12,0.22] | 0.580 | 401 | 0.06<br>[-0.11, 0.23] | 0.470 | 373 |

*Coefficient interpretation: average change in PCMC out of 90 points associated with attending a facility that had a labor & delivery readiness score of 1 percentage point higher*
\* significant at $p < 0.05$
<sup>+</sup> Lusaka model adjusted by 7 factors: women's employment, type of provider assisting during delivery, maternal PNC check received before discharge, PNC counseling on danger signs received before discharge, PNC counseling on family planning received before discharge, blood pressure checked before discharge, newborn PNC check received before discharge.
<sup>++</sup> Nairobi model adjusted by 10 factors: delivery facility type & managing authority, women's age, women's marital status, type of provider assisting during delivery, maternal PNC check received before discharge, PNC counseling on danger signs received before discharge, PNC counseling on family planning received before discharge, blood pressure checked before discharge, newborn PNC check received before discharge, appointment received for next PNC check before discharge.
<sup>+++</sup> Ouagadougou model adjusted by 11 factors: delivery facility type & managing authority, women's employment, women's age, ANC4+, type of provider assisting during delivery, maternal PNC check received before discharge, PNC counseling on danger signs received before discharge, PNC counseling on family planning received before discharge, blood pressure checked before discharge, newborn PNC check received before discharge, appointment received for next PNC check before discharge.

## Discussion

Our study in facilities serving informal settlements in Nairobi, Lusaka and Ouagadougou revealed sub-standard readiness for labor and delivery care, and even poorer readiness for SSNC. Readiness scores in informal settlement facilities ranged from 55.9% in Ouagadougou to 73.6% in Lusaka for labor and delivery care, and from 37.2% in Ouagadougou to 61.3% in Nairobi for SSNC. Importantly, we found that facilities serving informal settlements were statistically significantly less ready for both these services than those not serving informal settlements, although key items were lacking across all facilities. Our exact match linking analysis did not detect a statistically significant effect of higher labor and delivery readiness on women’s overall experience of PCMC.

Other studies have assessed readiness for MNH services in sub-Saharan Africa:(26,33–35) one study measuring childbirth care readiness in Ethiopia reported median scores ranging from 75% to 96%;(26) another using HHFA data in Malawi, Mozambique and Tanzania estimated SSNC readiness scores ranging from 53.7% to 80.7%.(36) A recent analysis in Burkina Faso highlighted a declining trend in the availability and readiness for emergency obstetric and newborn care between 2014 and 2018, driven by insufficient trained staff and medicines.

Authors suggested this may be linked to rising insecurity and the increased demand for health services since the adoption of free maternal health care in 2016.(35) While differences in readiness item selection and definition make comparisons across studies difficult, consistent with our findings, these studies also reported higher facility readiness among hospitals compared to health centers.(26,35)

Despite targeted efforts, substantial gaps in readiness for critical maternal and newborn care interventions persist across settings, and particularly in provider training. In our study, less than 8% and 30% of facilities serving and not serving informal settlements in Ouagadougou had staff trained in KMC and newborn resuscitation in the previous 2 years, respectively. In line with our findings, studies from Ethiopia and Northeastern Nigeria have highlighted health worker training gaps in the management of eclampsia and newborn resuscitation.(37,38) Importantly, there has been a lack of measures assessing provider knowledge and competency as health facility assessments do not measure the training content nor assess providers’ knowledge, posing an additional measurement constraint.(28)

KMC services offer the potential to save lives of preterm and low birthweight (LBW) babies(39) for whom skin-to-skin contact offers an inexpensive alternative to infant incubators, particularly in low-resource settings,(40,41) however substantial gaps persist. We found poor availability and readiness for KMC services across facilities serving and not serving informal settlements, especially in Ouagadougou. A systematic review of KMC implementation in SSA identified health systems barriers linked to insufficient space to carry out KMC, as well as inadequate staff training and guidelines.(42) This is in line with our results and others’(36): in Ouagadougou, we found that zero and 2.4% of facilities serving and not serving informal settlements respectively had a dedicated location for the caregiver to provide KMC overnight. Similarly, no facility serving informal settlements in Nairobi and Ouagadougou had KMC caps/hats for newborns. One of the factors explaining this result is that very few facilities offer KMC in Ouagadougou, and most newborns requiring KMC are referred to higher-level facilities, although there is no formal policy restricting the service to these facilities. Strong leadership prioritizing KMC implementation, readiness and provider training, especially in lower-level facilities serving vulnerable populations are critical to improve both access to and quality of SSNC, and ensure newborn survival.(42)

Addressing disparities in service readiness between facility types and managing authorities is essential in informal settlements where lower-level private facilities often lack adequate resources and regulation. Our analysis highlighted such differences: service readiness was higher in public hospitals serving informal settlements compared to those not serving informal settlements, and lowest in private facilities serving informal settlements, contrary to previous findings.(43) High delivery volumes in informal settlements may lead to more resources allocated to public hospitals in these areas, though further investigation is needed. Our findings also reflect the sprawling of unregulated, poorly equipped private facilities in low-income urban neighborhoods in SSA.(44,45) Efforts are needed to ensure that private facilities serving informal settlements are adequately regulated and ready to offer essential services. Importantly, the lower readiness in facilities serving informal settlement also reflects the combination of facility types in these areas, with fewer hospitals and more health centers, compared to facilities not serving informal settlements. For instance, in Ouagadougou, only two public hospitals served informal settlements, compared to ten outside informal settlements.

Consistent with our findings, several studies have reported higher readiness for MNH services in higher-level facilities.(28,46,47) An analysis in five African countries found that primary level facilities, which accounted for more than 90% of facilities offering obstetric services, had very low quality of maternal care based on readiness, referral systems and routine emergency care.(43) The latest DHS surveys suggest that in the Centre region of Burkina Faso, over 65% of births occurred in lower-level facilities (outside hospitals),(48) which also made up the majority of facilities serving informal settlements in our study, whereas in Nairobi county and Lusaka province, most births occurred in hospitals.(49,50) Quality improvement efforts are therefore critical in facilities where the majority of births occur.

While we hypothesized that facilities with better structural quality would offer more competent, empowered and motivated staff, and this would translate into a better experience of care, our findings did not support this. Other studies have reported significant but small magnitudes for the effect of structural quality on process quality of antenatal and sick child care in LMICs. (51,52) Most components of PCMC refer to women’s interactions with health providers, and their own perspectives on structural aspects such as the cleanliness, crowdedness and safety of the facility. These dimensions of PCMC are not directly captured in facility readiness indices, potentially explaining the lack of effect found. Importantly, facility readiness does not reflect health providers’ workload, stress/burnout, implicit bias and motivations, which have a large role in the provision of PCMC.(53–55) In line with our previous findings of poorer communication experiences in hospitals (compared to health centers), our supplementary analyses identified a small but significant negative effect of facility readiness on women’s communication and autonomy scores in Nairobi’s urban informal settlements. This reinforces the result that higher structural quality does not necessarily translate into health providers’ ability to offer person-centered care. Further research is needed to understand health providers’ barriers and enablers of PCMC in the context of urban informal settlements.

Our study has several strengths: to our knowledge, it is the first study comparing service readiness between facilities serving and not serving urban informal settlements residents, in addition to a multi-country comparison. The exact match linking between women’s client exit surveys and health facility assessments allowed for triangulation across data sources. Our results offer actionable insights for country-specific targeted interventions addressing the unavailability of key items needed for quality care. There are also limitations worth mentioning: facility readiness scores for labor and delivery and SSNC are influenced by the items that define them, and are not standardized across studies; item selection in our study was based on existing literature and WHO guidelines, and was identical for facilities serving and not serving informal settlements. PCMC scores relied on women’s self-report and were subject to recall bias; this was minimized by limiting the recall period to 2 weeks following facility discharge. Specific components of the PCMC scale are inherently subjective, and poor provider behavior may be normalized in low-resource settings.

## Conclusions

In conclusion, our study brings to light sub-standard service readiness for labor and delivery care and SSNC across facilities in Nairobi, Lusaka and Ouagadougou. Those serving urban informal settlements were particularly disadvantaged. While we found no significant association between labor and delivery care readiness and women’s overall experience of PCMC, further research is needed to better understand the relationship between structural quality, process quality, and experience of care. Despite nearly universal institutional deliveries in African cities, the road to meeting the Every Newborn Action Plan and Sustainable Development Goal targets for maternal and newborn health are still long. Health systems investments are warranted to improve the availability and functionality of key items, as well as ensure staff are trained in critical service areas, across facility types and both public and private sectors. Quality of care for essential MNH services remains inadequate for marginalized urban populations, and it is imperative to ensure that facilities serving these populations have the structural quality in place, both in terms of physical and human resources, to drive improvements in maternal and newborn survival.

## Data Availability

de-identified data analyzed in this study can be made available upon reasonable request to the corresponding author.

## Declarations

### Ethics approval and consent to participate

This study underwent ethical review and received approvals in each study site (AMREF approval P1309-2022and NACOSTI research license 686471 in Kenya, Comité d’éthique pour la Recherche en Santé approval 2022-10-216 in Burkina Faso, and University of Zambia Biomedical Research Ethics Committee approval 3215-2022 in Zambia), in addition to Johns Hopkins University IRB (approvals 22309 & 22304). All women who participated in client exit surveys provided written consent to participate.

## Availability of data and materials

The datasets generated and/or analysed during the current study are not publicly available as they belong to the institutions that collected them, but are available from the corresponding author on reasonable request.

## Acknowledgements

We are grateful to the facilities and their staff for agreeing to participate in our Nairobi study and in the HHFA assessment implemented by WHO in Lusaka and Ouagadougou. We are thankful to all the women who agreed to participate in the client exit surveys to share their experience of childbirth care. We thank and acknowledge the hard work of study team members in Nairobi, Lusaka and Ouagadougou in particular data collectors and field supervisors. We thank Mr. Ovost Chooye of Zambia’s Ministry of Health for support with facility mapping. We recognize Countdown to 2030 colleagues who contributed towards the study design, implementation, analysis and dissemination of findings.

## Authors’ contributions

SS Jiwani and A Amouzou conceptualized the study, with inputs from C Faye and T Boerma. SS Jiwani developed the study tools, with inputs and review from A Amouzou and A Sheffel. The study was implemented by SS Jiwani, M Mutua, C Jacobs, K Cisse, A Njeri, G Adero and M Musukuma. SS Jiwani led the statistical analysis and developed the first draft of the manuscript with inputs from A Amouzou. M Munos and E Stierman critically reviewed the findings. All co-authors reviewed and contributed to the final version of this manuscript.

## Consent for publication

not applicable

