## Supplementary materials for "Hidden Cities, Hidden Gaps: Measuring Facility Readiness for Maternal and Newborn Health Services and its Association with Person-Centered Maternity Care in Urban Informal Settlements of Nairobi, Lusaka and Ouagadougou cities"

Supplementary table 1. Definitions of items included in labor and delivery readiness index.

| Domain | Item description | Lusaka | Nairobi | Ouagadougou |
|---|---|---|---|---|
| Equipment | Blank partograph: observed available and reported functionality in service site | x | x | x |
| | Examination light: Functioning spotlight source that can be used for patient examinations; a functional flashlight is accepted. Observed available and reported functionality in service site | x | x | x |
| | blood pressure apparatus: Digital BP machine or manual sphygmomanometer with stethoscope. Observed available and reported functionality in service site. | x | x | x |
| | foetal stethoscope: Foetal stethoscope/pinard/ foetoscope/digital doplar. Observed and functional in service site | x | x | x |
| | delivery bed: delivery bed w/stirrups. Observed available and reported functional in service site | x | x | x |
| | Delivery pack (or cord clamp, episiotomy scissors, scissors/blade to cut cord, suture material with needle, and needle holder). Observed available and reported functionality in service site | x | x | x |
| | vacuum aspirator or D&C kit and speculum. Observed available and reported functionality in service area | x | x | x |
| | manual vacuum extractor or forceps for outlet application. Observed available and reported functionality in service area. | x | x | x |
| | sterilization equipment: dry heat sterilizer or an autoclave; if the machine is not electric, the heat source needs to be available and (If relevant) functioning (e.g., wood or gas is present for the autoclave). Observed available and reported functional anywhere in the facility | x | x | x |

| | | | | |
|---|---|---|---|---|
| | resuscitation table for newborn with heat source. Observed available and reported functional in service area. | x | x | x |
| | infant incubator. Observed available and reported functional anywhere in facility. | x | x | x |
| | suction pump and catheter or suction bulb for newborn. Observed available and reported functional in service area. | x | x | x |
| | neonatal resuscitation bag and mask (size 1 & size 0). Observed available and reported functionality in service site | x | x | x |
| | adult resuscitation bag and mask. Observed available and reported functionality in service site | x | x | x |
| | infant weighing scale with 100g gradation. Observed available and reported functionality in service site | x | x | x |
| | pulse oximeter. Observed available and reported functionality in service site | x | x | x |
| | thermometer: Manual, electronic or digital. Observed available and reported functionality in service site | x | x | x |
| | latex gloves: If equivalent non latex gloves are available this is acceptable; Either sterile or non sterile; latex or equivalent. Observed available in service site | x | x | x |
| | hand hygiene: soap and water, or alcohol based rub. Observed in service site. | x | x | x |
| | clean towel for drying newborn. Observed available in service site | x | x | x |
| | non-sharps waste container: Waste receptacle bin with lid and plastic bin liner for non-sharps infectious waste. Observed available in service site. | x | x | x |
| | sharps container. Observed available in service site. | x | x | x |
| | environmental disinfectant. Observed available in service site. | x | x | x |
| Medicines & commodities | Injectable uterotonic: Oxytocin (or country-specific equivalent) observed and stored in cold storage, at least one not expired in service site or where routinely stored. | x | x | x |
| | injectable betamethasone or dexamethasone. Observed at least one not expired in service site or where routinely stored | x | x | x |
| | IV solution and perfusion kit: Intravenous solution for infusing medicines (Ringers lactate, Normal saline, or D5W) and infusion set. Observed available and at least one not expired in service site or where routinely stored. | x | x | x |

| | | | | |
|---|---|---|---|---|
| | Misoprostol tablet 200 mcg. Observed at least one not expired in service site or where routinely stored | x | x | x |
| | oxygen and administration equipment: Oxygen currently in unit or oxygen currently not in unit, but report that unit calls centrally for O2 if needed and humidifier, flowmeter, oxygen delivery apparatus are observed available and functioning in service site. | x | x | x |
| | antibiotic eye ointment: Eye cream for newborn or for trachoma (tetracycline or country-specific equivalent). Observed at least one not expired where routinely stored | x | x | x |
| | injectable magnesium sulphate, 50% strength or alternative. Observed and not expired in service site or where routinely stored | x | x | x |
| | injectable antibiotic for maternal/neonatal sepsis: Broad-spectrum injectable antibiotic treatment of sepsis in mother and newborn. Specific combination- (Ampicillin powder for injection + gentamicin injection) or (penicillin injection + gentamicin injection) or ceftriaxone injection. Observed at least one not expired in service site or where routinely stored. | x | x | x |
| | skin disinfectant for cord care: 4% chlorhexidine.* Observed available and at least one not expired in service site or where routinely stored | | x | x |
| | injectable Vitamin K. observed available and not expired in service site or where routinely stored | x | x | x |
| | injectable calcium gluconate. observed available and not expired in service site or where routinely stored | x | x | x |
| | Hypertensive (hydralazine, nifedipine, or methyldopa). Observed at least one not expired in service site or where routinely stored | x | x | x |
| | injectable metronidazole. Observed at least one not expired in service site or where routinely stored | x | x | x |
| | injectable diazepam. Observed at least one not expired in service site or where routinely stored | x | | x |
| Human resources & guidelines | At least one staff in service area trained in newborn resuscitation using bag and mask in last 2 years | x | x | x |
| | At least one staff in service area trained in essential childbirth care in the last two years | x | x | x |

| | | | | |
|---|---|---|---|---|
| | At least one staff in service area trained in essential newborn care in last 2 years | x | x | x |
| | guidelines for essential childbirth care observed in service site. | x | x | x |
| | checklist or job aids for essential childbirth care observed in service site. | x | x | x |
| | guidelines for essential newborn care observed in service site. | x | x | x |
| | skilled attendant present on call (onsite or in near proximity) 24 hours | x | x | x |
| Basic amenities | improved water source: Main water source located on premises and from which water is available. Improved water sources include: piped, public tap, standpipe, tubewell/borehole, protected dug well, protected spring, rain water. If public tap, standpipe, tubewell/borehole, protected dug well, protected spring, rain water. | x | x | x |
| | improved sanitation facility: Flush/pour flush to piped sewer system or septic tank, pit latrine with slab, or composting toilet available in service site or oupatient area. | x | x | x |
| | electricity available at all times when facility was open in last 7 days and/or back-up energy | x | x | x |
| | emergency transport (available on day of survey): Facility has access to a functional ambulance or other vehicle for emergency transportation for clients that is either stationed at this facility or that the facility can call for, and vehicle available and functional today | x | x | x |
| Performance of BeMONC signal functions in last 12 months | Parenteral administration of antibiotics (IV or IM) for mothers | x | x | x |
| | Parenteral administration of anticonvulsants (magnesium sulfate) for management of pre-eclampsia and eclampsia (IV or IM) | x | x | x |
| | Parenteral administration of oxytocic for treatment of postpartum hemorrhage (IV or IM) | x | x | x |
| | Removal of retained products of conception using D&C or manual vacuum aspiration | x | x | x |
| | Assisted vaginal delivery using manual vacuum extraction (MVE) or forceps | x | x | x |
| | Manual removal of placenta | x | x | x |
| | Neonatal resuscitation with bag and mask | x | x | x |

*based on country-specific recommendations. Chlorhexidine for cleaning cord excluded in Lusaka as it is not a national policy.

Supplementary table 2. Definitions of items included in small & sick newborn care readiness index.

| Domain | item description | Lusaka | Nairobi | Ouagadougou |
|---|---|---|---|---|
| **Immediate newborn care and routine care** | clean towel for drying newborn. Observed available in service site | x | x | x |
| | cord cutting supplies: sterile scissors and/or sterile blade to cut cord AND umbilical cord clamp (sterile ligatures or clamp of Barr) OR delivery pack. Observed available and functional in service site. | x | x | x |
| | radiant heater/warmth source or infant incubator. Observed available and reported functionality anywhere in the facility. | x | x | x |
| | Injectable Vitamin K. observed available and not expired in service site or where routinely stored. | x | x | x |
| | antibiotic eye ointment: Eye cream for newborn or for trachoma (tetracycline or country-specific equivalent). Observed at least one not expired where routinely stored | x | x | x |
| | skin disinfectant for cord care: 4% chlorhexidine.* Observed available and at least one not expired in service site or where routinely stored | | x | x |
| | immunization supplies: 1) BCG, 2) Hep B, and 3) OPV observed in pharmacy or anywhere in the facility where medicines are routinely stored; at least one with valid expiration date observed available. 4) Single use needles and syringes are observed available in the delivery service provision area, OPD, pharmacy, or immunization service area.<br>5) Guidelines containing information on child immunization or the national immunization schedule are observed available in the immunization service provision area.<br>6) A refrigerator dedicated for vaccine storage with a temperature monitoring device and consistent electricity supply is available in the facility OR the facility has a cold box/vaccine carrier with ice packs in the immunization service area. Calculated as proportion of items available out of 6. | x | x | x |
| **Early initiation and support for breastfeeding** | Guidelines for breastfeeding and promotion of breastfeeding. Observed available in service site. | x | x | x |
| | At least one staff in service site trained in breastfeeding and promotion of breastfeeding in last two years | x | x | x |

| | | | | |
|---|---|---|---|---|
| | Facility routinely practices breastfeeding initiation within 1 hour of birth | x | x | x |
| **Neonatal resuscitation** | Airway suction apparatus: suction apparatus with catheter or suction bulb for mucus extraction. Observed available and reported functionality in service site. | x | x | x |
| | neonatal resuscitation bag and mask (size 1 & size 0). Observed available and reported functionality in service site | x | x | x |
| | resuscitation table for newborn with heat source. Observed available and reported functional in service area. | x | x | x |
| | At least one staff in service area trained in newborn resuscitation using bag and mask in last 2 years | x | x | x |
| | Facility provided neonatal resuscitation in past 12 months. | x | x | x |
| **Prevention of mother-to-child transmission of HIV (PMTCT)** | PMTCT room is private room with auditory and visual privacy.** | x | x | x |
| | Antiretrovirals for newborns: nevirapine syrup or zidovudine syrup. Observed available and at least one unexpired in service site or where routinely stored. | x | x | x |
| | Antiretrovirals for mothers: country-specific first line ARV prophylaxis for HIV-positive pregnant women. Observed available and at least one unexpired in service site or where routinely stored. | x | x | x |
| | Cotrimoxazole syrup. Observed available and at least one unexpired in service site or where routinely stored. | x | | x |
| | HIV diagnostic capacity: facility has the ability to conduct quantitative nucleic acid testing for HIV diagnosis onsite (PCR) or RNA / PCR for DNA-EID and has filter paper for DBS, or ELISA test with ELISA washer, ELISA reader, incubator, specific assay kit available and functional; Or facility sends specimen offsite for testing. | x | x | x |
| | PMTCT and infant and young child feeding (IYCF) guidelines available observed in service site. Calculated as proportion of guidelines available out of 2. | x | | x |
| | At least one staff in service site trained in PMTCT in last two years. | x | | x |
| | At least one staff in service site trained in newborn nutrition counseling of mother with HIV or IYCF in last 2 years. | x | | x |
| **Kangaroo Mother Care (KMC)** | bed/location for caregiver to stay overnight to provide KMC observed. | x | x | x |
| | caps/hats for newborn observed available. | x | x | x |
| | Register to record KMC services observed available. | x | x | |

| | | | | |
|---|---|---|---|---|
| | Guidelines, protocols or job aids for KMC observed available. | x | x | x |
| | At least one staff in service site trained in KMC in last two years. | x | x | x |
| | Facility provided KMC in last 3 months. | x | x | |
| **Detection and management of neonatal infection** | Antibiotic treatment for neonatal infection (Outpatient treatment: Gentamycin (IM or IV) and Amoxicillin (syrup/oral suspension) OR Inpatient treatment: (Benzylpenicillin (IV or IM) OR Ampicillin (IV or IM)) AND Gentamycin (IM or IV)) observed in pharmacy or anywhere in the facility where medicines are routinely stored; at least one with valid expiration date observed. | x | x | x |
| | full blood count diagnostic capacity: Facility has the ability to conduct full blood count onsite and has functioning hematology analyzer or functioning blood chemistry analyzer available and functioning; or facility sends specimen offsite for testing. | x | x | x |
| | chest x-ray capacity: Facility has the ability to conduct diagnostic x-ray testing + digital x-ray machine (not requiring film with equipment available) or x-ray machine and unexpired film and equipment available and functioning. | x | | x |
| | register for neonatal sepsis observed in service site. | x | x | x |
| | At least one staff in service site trained in neonatal sepsis in last two years. | x | x | x |
| | guidelines/checklists for neonatal sepsis observed available in service site. | x | x | x |
| **Comfort and pain management** | Paracetamol syrup observed and at least one unexpired in service site or where routinely stored. | x | | x |
| | morphine (syrup or injectable) observed and at least one unexpired in service site or where routinely stored. | x | | x |
| **Detection and management of hypoglycemia** | Glucose injectable solution (30% or 50%) observed and at least one unexpired in service site or where routinely stored. | x | | x |
| | Blood glucose testing capacity: Facility has the ability to conduct blood glucose testing onsite and has glucometer and test strips available and functioning or blood chemistry analyzer available and functioning; or sends blood chemistry specimen offsite for testing. | x | | x |
| **General facility readiness** | Thermometer: observed available and functional in service site | x | x | x |
| | infant weighing scale with 100g gradation. Observed available and reported functionality in service site | x | x | x |

| | | | | |
|---|---|---|---|---|
| | stethoscope. Observed available and reported functionality in service site | x | x | x |
| | medication delivery mechanism: pediatric infusion kit + any IV fluid observed anywhere in the facility where supplies are routinely stored OR single use syringes observed in the delivery service provision area | x | x | x |
| | pulse oximeter. Observed available and reported functionality in service site | x | x | x |
| | oxygen and administration equipment: Oxygen currently in unit or oxygen currently not in unit, but report that unit calls centrally for O2 if needed and humidifier, flowmeter, oxygen delivery apparatus are observed available and functioning in service site. | x | x | x |
| | non-sharps waste container: Waste receptacle bin with lid and plastic bin liner for non-sharps infectious waste. Observed available in service site. | x | x | x |
| | sharps container. Observed available in service site. | x | x | x |
| | environmental disinfectant. Observed available in service site. | x | x | x |
| | latex gloves: If equivalent non latex gloves are available this is acceptable; Either sterile or non sterile; latex or equivalent. Observed available in service site | x | x | x |
| | hand hygiene: soap and water, or alcohol based rub. Observed in service site. | x | x | x |
| | guidelines for essential newborn care or referral of small or sick newborn observed available in service site. | x | x | x |

*based on country-specific recommendations. Chlorhexidine for cord care excluded in Lusaka as it is not a national policy.

**In Nairobi, used outpatient room with auditory and visual privacy instead of PMTCT room, due to data unavailability.

Supplementary table 3. PCMC scale questions, response categories and scoring

| Question | Response categories | PCMC scoring (out of 90) |
|---|---|---|
| **DIGNITY & RESPECT** | | |
| Did the doctors, nurses, or other staff at the facility treat you with respect? | No, never | 0 |
| | Yes, a few times | 1 |
| | Yes, most of the time | 2 |
| | Yes, all the time | 3 |
| Did the doctors, nurses, or other staff at the facility treat you in a friendly manner? | No, never | 0 |
| | Yes, a few times | 1 |
| | Yes, most of the time | 2 |
| | Yes, all the time | 3 |
| Did you feel that they shouted at you, scolded, insulted, threatened, or talked to you rudely? | No, never | 3 |
| | Yes, once | 2 |
| | Yes, a few times | 1 |
| | Yes, many times | 0 |
| | Refused to respond | 3 |
| Did you feel like you were treated roughly like pushed, beaten, slapped, pinched, physically restrained, or gagged? | No, never | 3 |
| | Yes, once | 2 |
| | Yes, a few times | 1 |
| | Yes, many times | 0 |
| | Refused to respond | 3 |
| During examinations in the labor room, were you covered up with a cloth or blanket, or screened with a curtain so that you did not feel exposed? | No, never | 0 |
| | Yes, a few times | 1 |
| | Yes, most of the time | 2 |
| | Yes, all the time | 3 |
| Do you feel like your health information was or will be kept confidential at this facility? | No, never | 0 |
| | Yes, a few times | 1 |
| | Yes, most of the time | 2 |

| | | |
|---|---|---|
| | Yes, all the time | 3 |
| **COMMUNICATION & AUTONOMY** | | |
| During your time in the health facility did the doctors, nurses, or other health-care providers introduce themselves to you when they first came to see you? | No, none of them | 0 |
| | Yes, a few of them | 1 |
| | Yes, most of them | 2 |
| | Yes, all of them | 3 |
| Did the doctors, nurses, or other health-care providers call you by your name? | No, never | 0 |
| | Yes, a few times | 1 |
| | Yes, most of the time | 2 |
| | Yes, all the time | 3 |
| Did you feel like the doctors, nurses or other staff at the facility involved you in decisions about your care? | No, never | 0 |
| | Yes, a few times | 1 |
| | Yes, most of the time | 2 |
| | Yes, all the time | 3 |
| | Did not have to make any decisions | 3 |
| During the delivery, do you feel like you were able to be in the position of your choice? | No, never | 0 |
| | Yes, for a short time | 1 |
| | Yes, most of the time | 2 |
| | Yes, all the time | 3 |
| Did the doctors, nurses, or other staff at the facility speak to you in a language you could understand? | No, never | 0 |
| | Yes, a few times | 1 |
| | Yes, most of the time | 2 |
| | Yes, all the time | 3 |
| Did the doctors, nurses, or other staff at the facility ask your permission or consent before doing procedures on you? | No, never | 0 |
| | Yes, a few times | 1 |
| | Yes, most of the time | 2 |
| | Yes, all the time | 3 |
| Did the doctors, nurses, or other staff at the facility explain to you why they were doing examinations or procedures on you? | No, never | 0 |
| | Yes, a few times | 1 |
| | Yes, most of the time | 2 |

| | | |
|---|---|---|
| | Yes, all the time | 3 |
| Did the doctors, nurses, or other staff at the facility explain to you why they were giving you any medicine? | No, never | 0 |
| | Yes, a few times | 1 |
| | Yes, most of the time | 2 |
| | Yes, all the time | 3 |
| | Did not get any medicine | 3 |
| Did you feel you could ask the doctors, nurses, or other staff at the facility any questions you had? | No, never | 0 |
| | Yes, a few times | 1 |
| | Yes, most of the time | 2 |
| | Yes, all the time | 3 |
| **SUPPORTIVE CARE** | | |
| How did you feel about the amount of time you waited to receive care? Would you say it was: | Very short | 3 |
| | Somewhat short | 2 |
| | Somewhat long | 1 |
| | Very long | 0 |
| Did the doctors and nurses at the facility show concern for your feelings about your delivery? | No, never | 0 |
| | Yes, a few times | 1 |
| | Yes, most of the time | 2 |
| | Yes, all the time | 3 |
| Did the doctors, nurses, or other staff at the facility try to understand your anxieties? | No, never | 0 |
| | Yes, a few times | 1 |
| | Yes, most of the time | 2 |
| | Yes, all the time | 3 |
| | Did not have any anxiety | 3 |
| When you needed help, did you feel the doctors, nurses, or other staff at the facility paid attention? | No, never | 0 |
| | Yes, a few times | 1 |
| | Yes, most of the time | 2 |
| | Yes, all the time | 3 |
| Do you feel the doctors or nurses did everything they could to help control your pain? | No, never | 0 |
| | Yes, a few times | 1 |
| | Yes, most of the time | 2 |

| | | |
|---|---|---|
| | Yes, all the time | 3 |
| Were you allowed to have someone you wanted (outside of staff at the facility, such as family or friends) to stay with you during labor? | No, never | 0 |
| | Yes, a few times | 1 |
| | Yes, most of the time | 2 |
| | Yes, all the time | 3 |
| | I did not want someone to stay with me | 3 |
| Were you allowed to have someone you wanted to stay with you during delivery? | No, never | 0 |
| | Yes, a few times | 1 |
| | Yes, most of the time | 2 |
| | Yes, all the time | 3 |
| | I did not want someone to stay with me | 3 |
| Did you feel the doctors, nurses, or other staff at the facility took good care of you, at the best of their ability? | No, never | 0 |
| | Yes, a few times | 1 |
| | Yes, most of the time | 2 |
| | Yes, all the time | 3 |
| Did you feel you could completely trust the doctors, nurses, or other staff at the facility with regards to your care? | No, never | 0 |
| | Yes, a few times | 1 |
| | Yes, most of the time | 2 |
| | Yes, all the time | 3 |
| Do you think there were enough health staff in the facility to care for you? | No, never | 0 |
| | Yes, a few times | 1 |
| | Yes, most of the time | 2 |
| | Yes, all the time | 3 |
| Thinking about the labor and postnatal wards, did you feel the health facility was crowded? | No, never | 3 |
| | Yes, a few times | 2 |
| | Yes, most of the time | 1 |
| | Yes, all the time | 0 |
| Thinking about the wards, washrooms, and the general environment of the health facility, would you | Very dirty | 0 |
| | Dirty | 1 |

| | | |
|---|---|---|
| say the facility was very clean, clean, dirty, or very dirty? | Clean | 2 |
| | Very clean | 3 |
| Was there running water in the facility? | No, never | 0 |
| | Yes, a few times | 1 |
| | Yes, most of the time | 2 |
| | Yes, all the time | 3 |
| Was there electricity in the facility? | No, never | 0 |
| | Yes, a few times | 1 |
| | Yes, most of the time | 2 |
| | Yes, all the time | 3 |
| In general, did you feel safe in the health facility? | No, never | 0 |
| | Yes, a few times | 1 |
| | Yes, most of the time | 2 |
| | Yes, all the time | 3 |

Supplementary table 4. Service availability in facilities offering maternal and newborn health services by city.

| | LUSAKA | | | | NAIROBI | | OUAGADOUGOU | | | |
|---|---|---|---|---|---|---|---|---|---|---|
| | Serving any informal settlement | | Not serving informal settlements | | Serving informal settlements | | Serving informal settlements | | Not serving informal settlements | |
| | (%) | n | (%) | n | (%) | n | (%) | n | (%) | n |
| **Labor & Delivery Care** | 35.5 | 31 | 30.7 | 88 | 47.4 | 38 | 47.0 | 115 | 39.1 | 215 |
| BeMONC (facility reported) | 32.3 | 31 | 29.6 | 88 | 47.4 | 38 | 41.7 | 115 | 36.3 | 215 |
| Parenteral administration of antibiotics (IV or IM) for mothers | 32.3 | 31 | 29.6 | 88 | 47.4 | 38 | 44.3 | 115 | 38.1 | 215 |
| Parenteral administration of anticonvulsants (magnesium sulfate) for management of pre-eclampsia and eclampsia (IV or IM) | 32.3 | 31 | 29.6 | 88 | 42.1 | 38 | 18.3 | 115 | 19.1 | 215 |
| Parenteral administration of oxytocic for treatment of postpartum haemorrhage (IV or IM) | 32.3 | 31 | 30.7 | 88 | 42.1 | 38 | 44.3 | 115 | 37.7 | 215 |
| Removal of retained products of conception using D&C or manual vacuum aspiration | 16.1 | 31 | 25.0 | 88 | 31.6 | 38 | 33.9 | 115 | 24.7 | 215 |
| Assisted vaginal delivery using manual vacuum extraction (MVE) or forceps | 12.9 | 31 | 20.5 | 88 | 23.7 | 38 | 29.6 | 115 | 23.7 | 215 |
| Manual removal of placenta | 32.3 | 31 | 30.7 | 88 | 47.4 | 38 | 44.3 | 115 | 35.8 | 215 |
| Neonatal resuscitation with bag and mask | 32.3 | 31 | 30.7 | 88 | 44.7 | 38 | 21.7 | 115 | 17.7 | 215 |
| **All 7 BeMONC signal functions** | 9.7 | 31 | 20.5 | 88 | 15.8 | 38 | 9.6 | 115 | 4.7 | 215 |
| Oxygen administration | 32.3 | 31 | 30.7 | 88 | 39.5 | 38 | 5.2 | 115 | 9.3 | 215 |
| CeMONC (among hospitals, facility reported) | 100.0 | 4 | 77.8 | 9 | 66.7 | 3 | 23.5 | 17 | 27.8 | 72 |
| Blood transfusion (among hospitals) | 75.0 | 4 | 55.6 | 9 | 66.7 | 3 | 23.5 | 17 | 22.2 | 72 |
| C section (among hospitals) | 75.0 | 4 | 55.6 | 9 | 66.7 | 3 | 23.5 | 17 | 25.0 | 72 |
| **Maternal And Well Infant Postpartum/ Postnatal Care (inpatient, outpatient or both)*** | 71.0 | 31 | 62.5 | 88 | 68.4 | 38 | 49.6 | 115 | 42.3 | 215 |
| Counseling on family planning | 64.5 | 31 | 58.0 | 88 | 65.8 | 38 | 47.0 | 115 | 42.3 | 215 |
| Provision of newborn vaccines (BCG & OPV0) | 54.8 | 31 | 45.5 | 88 | 55.3 | 38 | 37.4 | 115 | 29.8 | 215 |

| | | | | | | | | | | |
|---|---|---|---|---|---|---|---|---|---|---|
| Counselling on child nutritional needs and good feeding practices | 64.5 | 31 | 55.7 | 88 | 63.2 | 38 | 48.7 | 115 | 41.4 | 215 |
| Counselling on danger signs in the newborn | 64.5 | 31 | 56.8 | 88 | 65.8 | 38 | 49.6 | 115 | 42.3 | 215 |
| Counselling on cord care and hygiene | 61.3 | 31 | 56.8 | 88 | 63.2 | 38 | 49.6 | 115 | 42.3 | 215 |
| Counselling on child immunization needs | 64.5 | 31 | 53.4 | 88 | 63.2 | 38 | 48.7 | 115 | 40.9 | 215 |
| **Small or Sick Newborn Care** | 54.8 | 31 | 43.2 | 88 | 86.8 | 38 | 13.0 | 115 | 20.0 | 215 |
| Kangaroo Mother Care | 16.1 | 31 | 18.2 | 88 | 10.5 | 38 | 8.7 | 115 | 12.6 | 215 |
| Services or referral for cases of neonatal sepsis | 38.7 | 31 | 29.6 | 88 | 73.7 | 38 | 12.2 | 115 | 17.2 | 215 |
| Neonatal resuscitation | 32.3 | 31 | 30.7 | 88 | 44.7 | 38 | 23.5 | 115 | 20.0 | 215 |
| Prevention of mother-to-child transmission of HIV | 35.5 | 31 | 27.3 | 88 | 42.1 | 38 | 37.4 | 115 | 31.6 | 215 |
| Early initiation and support for breastfeeding | 35.5 | 31 | 30.7 | 88 | 47.4 | 38 | 47.0 | 115 | 38.6 | 215 |

**routinely provided to all clients*

Supplementary figure 1: Distribution of labor & delivery readiness scores comparing facilities serving and not serving informal settlements, by city.

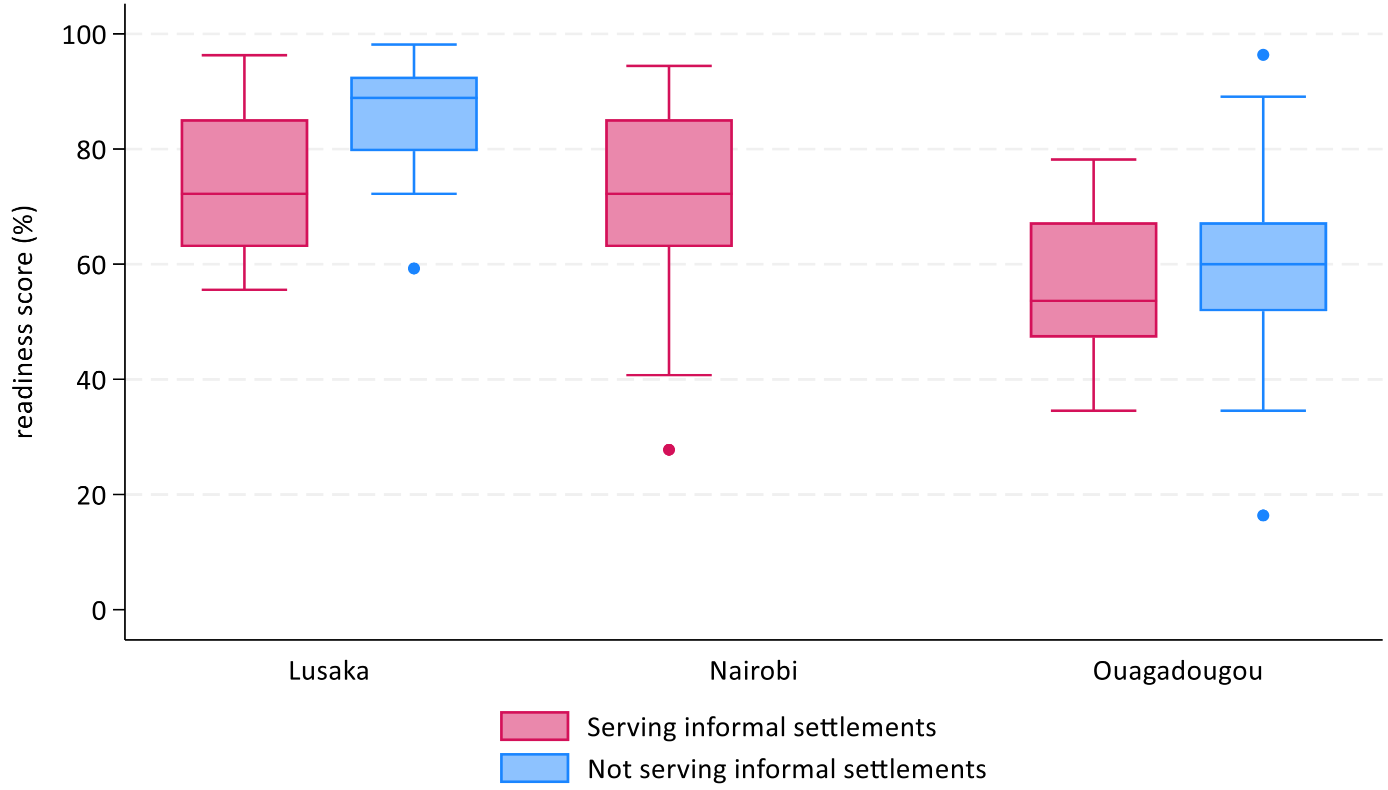

Supplementary table 5. Facility readiness scores (%) among facilities offering labor and delivery care by city.

| | LUSAKA | | | NAIROBI | OUAGADOUGOU | | |
|---|---|---|---|---|---|---|---|
| Domain | Serving informal settlements (n=11) % (SD) | Not serving informal settlements (n=27) % (SD) | t test p value | Serving informal settlements (n=18) % (SD) | Serving informal settlements (n=54) % (SD) | Not serving informal settlements (n=84) % (SD) | t test p value |
| ***Labor & delivery care*** | | | | | | | |
| Overall labor & delivery readiness *(simple additive score)* | 73.6 (12.5) | 85.6 (9.2) | **0.002** | 69.5 (19.4) | 55.9 (11.9) | 60.3 (12.7) | **0.044** |
| Equipment | 77.9 (12.7) | 93.2 (9.4) | **<0.001** | 65.7 (23.8) | 61.1 (13.6) | 65.7 (13.0) | 0.050 |
| Medicines & commodities | 61.5 (17.9) | 84.3 (12.9) | **<0.001** | 72.2 (23.2) | 46.7 (17.9) | 52.0 (22.1) | 0.145 |
| Basic amenities | 79.5 (18.8) | 90.7 (12.3) | **0.036** | 77.8 (19.0) | 70.8 (18.7) | 72.0 (17.5) | 0.705 |
| Human resources & guidelines | 75.3 (26.4) | 57.1 (24.7) | 0.051 | 68.3 (15.1) | 32.5 (25.0) | 40.8 (26.4) | 0.069 |
| Performance of BeMONC signal function in last 12 months | 76.6 (19.5) | 88.4% (20.2) | 0.110 | 73.8 (21.5) | 72.0 (23.2) | 71.9 (19.0) | 0.996 |
| *% facilities with score ≥95% (ceiling)* | *9.1* | *7.4* | | *0.0* | *0.0* | *1.2* | |
| *% facilities with score ≤5% (floor)* | *0.0* | *0.0* | | *0.0* | *0.0* | *0.0* | |
| ***Small & sick newborn care*** | | | | | | | |
| Overall SSNC readiness *(weighted additive score)* | 60.8 (14.5) | 70.7 (11.4) | **0.032** | 61.3 (17.9) | 37.2 (9.3) | 40.3 (12.4) | **0.010** |
| Immediate newborn care and routine care | 54.8 (18.4) | 82.2 (20.0) | **<0.001** | 66.3 (25.7) | 52.0 (18.2) | 55.0 (18.8) | 0.366 |
| Early initiation and support for breastfeeding | 81.8 (27.3) | 60.5 (29.3) | **0.045** | 74.1 (21.6) | 55.6 (25.9) | 61.5 (27.6) | 0.208 |

| | | | | | | | |
|---|---|---|---|---|---|---|---|
| Neonatal resuscitation | 80.0 (31.0) | 90.4 (12.9) | 0.305 | 77.8 (25.6) | 35.2 (22.0) | 40.5 (24.0) | 0.194 |
| Prevention of mother-to-child transmission of HIV | 85.8 (19.0) | 70.6 (21.4) | **0.047** | 77.8 (28.3) | 41.8 (18.7) | 43.5 (19.6) | 0.620 |
| Kangaroo Mother Care | 33.3 (34.2) | 27.2 (32.1) | 0.601 | 14.8 (29.1) | 1.9 (6.6) | 3.9 (14.3) | 0.264 |
| Detection and management of neonatal infection | 43.9 (25.0) | 51.9 (21.4) | 0.331 | 42.2 (23.7) | 20.7 (11.6) | 24.8 (19.6) | 0.124 |
| Comfort and pain management | 13.6 (23.4) | 66.7 (34.0) | **<0.001** | NA | 38.9 (21.0) | 35.7 (26.4) | 0.458 |
| Detection and management of hypoglycemia | 63.6 (39.3) | 94.4 (16.0) | **0.028** | NA | 16.7 (27.5) | 20.2 (31.2) | 0.493 |
| General facility readiness | 90.2 (9.7) | 92.3 (8.0) | 0.488 | 75.9 (20.8) | 72.5 (10.4) | 77.8 (10.0) | **0.004** |
| *% facilities with score ≥95% (ceiling)* | *0.0* | *0.0* | | *0.0* | *0.0* | *0.0* | |
| *% facilities with score ≤5% (floor)* | *0.0* | *0.0* | | *0.0* | *0.0* | *0.0* | |

Supplementary figure 2 A-C. Radar plots of labor & delivery care item availability by city.

A. Lusaka: facilities serving informal settlements (n=11)

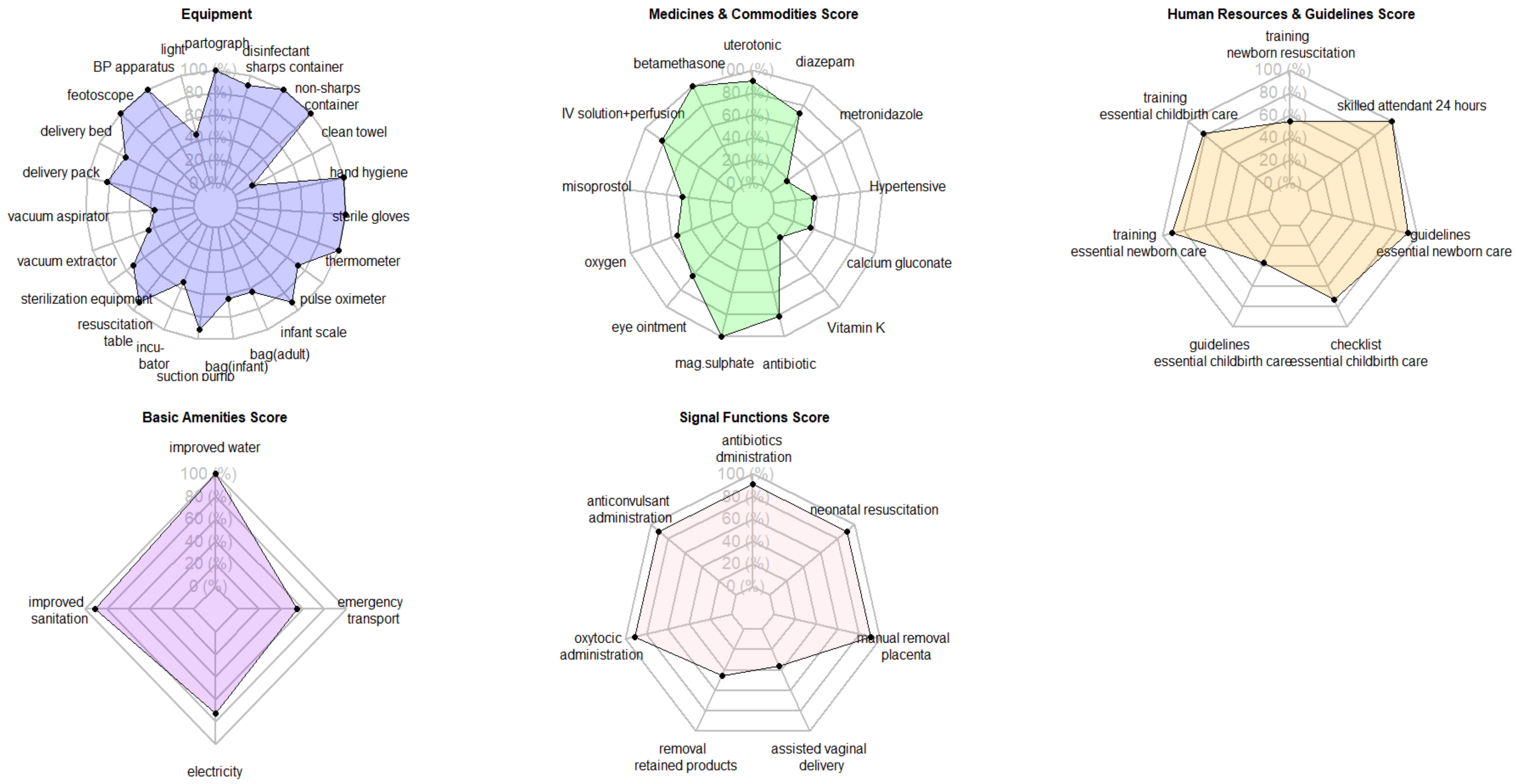

B. Nairobi: facilities serving informal settlements (n=18)

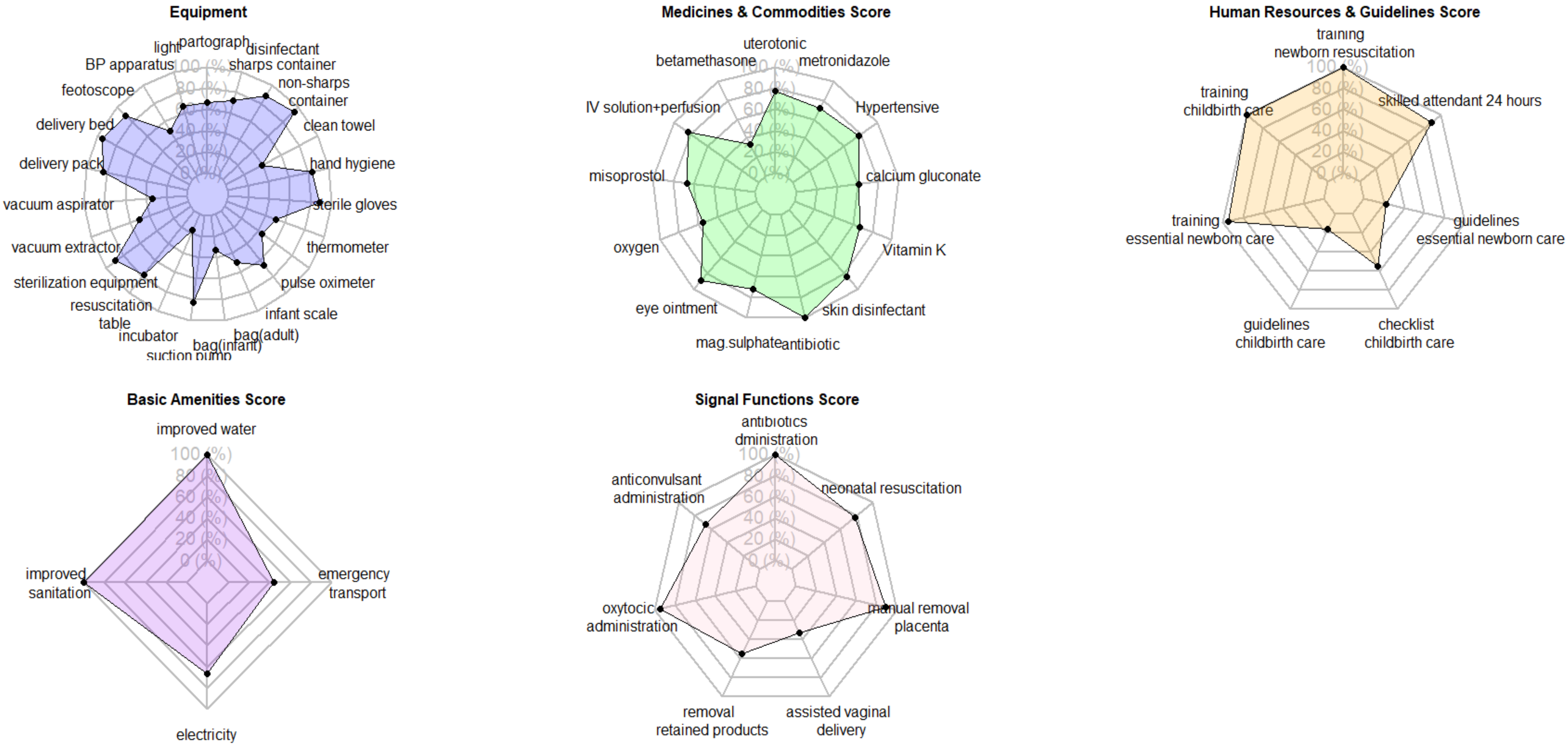

C. <u>Ouagadougou: facilities serving informal settlements (n=54)</u>

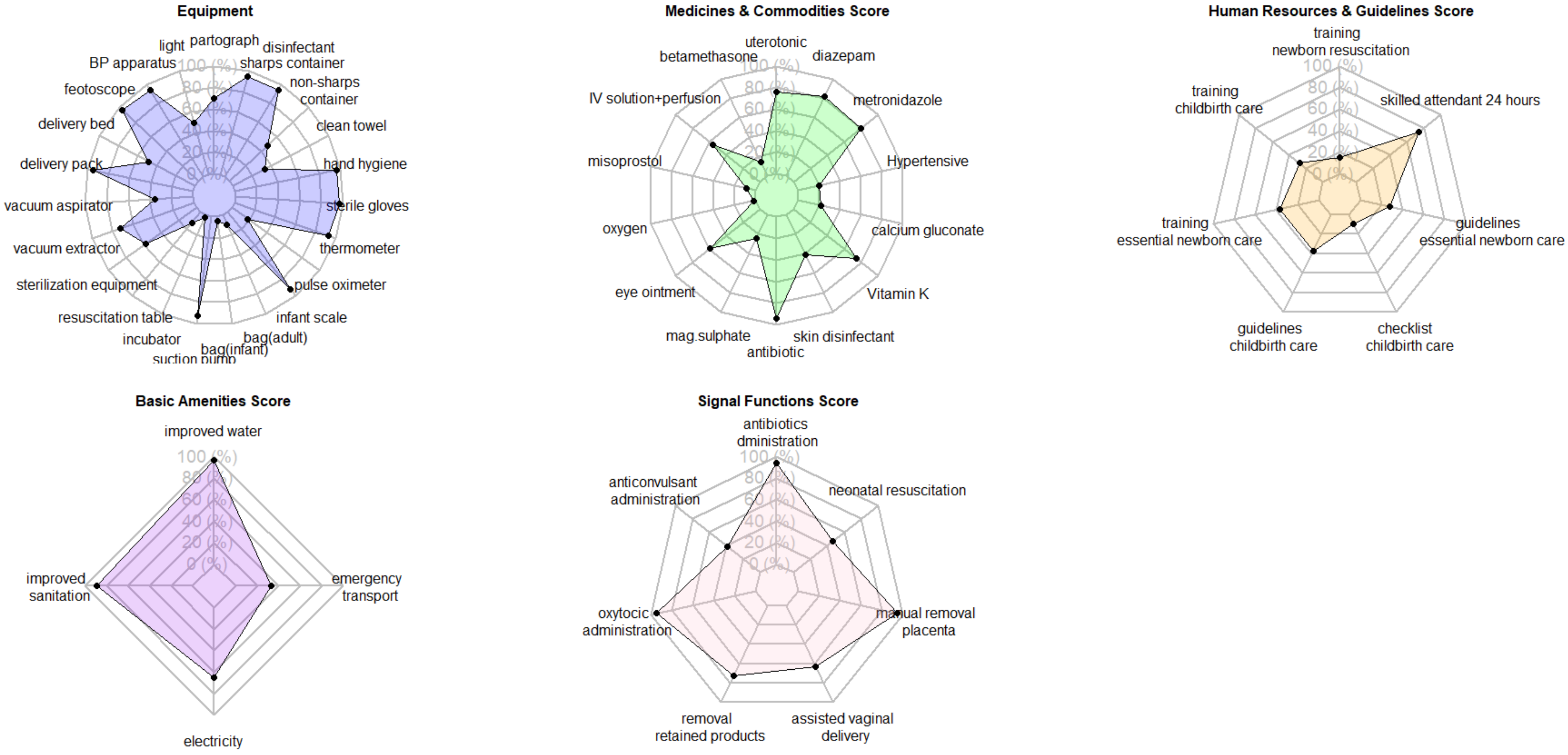

Supplementary table 6. Item availability for labor & delivery care readiness by city and facilities serving/not serving informal settlements.

| | | LUSAKA | | NAIROBI | OUAGADOUGOU | |
|---|---|---|---|---|---|---|
| Domain | Item | Serving informal settlements (n=11) | Not serving informal settlements (n=27) | Serving informal settlements (n=18) | Serving informal settlements (n=54) | Not serving informal settlements (n=84) |
| Equipment | Blank partograph | 100.0% | 96.3% | 66.7% | 70.4% | 71.4% |
| | examination light | 45.5% | 92.6% | 66.7% | 50.0% | 61.9% |
| | blood pressure apparatus | 100.0% | 100.0% | 50.0% | 94.4% | 95.2% |
| | foetal stethoscope | 100.0% | 100.0% | 88.9% | 96.3% | 97.6% |
| | delivery bed | 72.7% | 96.3% | 94.4% | 48.1% | 60.7% |
| | Delivery pack (or cord clamp, episiotomy scissors, scissors/blade to cut cord, suture material with needle, and needle holder) | 81.8% | 100.0% | 83.3% | 94.4% | 90.5% |
| | vacuum aspirator or D&C kit and speculum | 36.4% | 74.1% | 33.3% | 35.2% | 36.9% |
| | manual vacuum extractor or forceps | 45.5% | 85.2% | 50.0% | 72.2% | 81.0% |
| | sterilization equipment | 72.7% | 77.8% | 88.9% | 57.4% | 69.0% |
| | resuscitation table with heat source | 90.9% | 100.0% | 77.8% | 13.0% | 22.6% |
| | infant incubator | 54.5% | 81.5% | 16.7% | 1.9% | 9.5% |
| | suction pump and catheter or suction bulb (newborn) | 90.9% | 96.3% | 83.3% | 92.6% | 91.7% |
| | neonatal resuscitation bag and mask (size 1 & size 0) | 63.6% | 88.9% | 33.3% | 3.7% | 7.1% |
| | adult resuscitation bag and mask | 63.6% | 85.2% | 50.0% | 9.3% | 14.3% |
| | infant weighing scale (100g gradation) | 90.9% | 96.3% | 66.7% | 92.6% | 92.9% |
| | pulse oximeter | 72.7% | 92.6% | 44.4% | 18.5% | 27.4% |
| | thermometer | 100.0% | 100.0% | 50.0% | 92.6% | 95.2% |
| | sterile gloves | 100.0% | 100.0% | 88.9% | 96.3% | 98.8% |
| | hand hygiene | 100.0% | 100.0% | 83.3% | 96.3% | 94.0% |
| | clean towel for drying newborn | 18.2% | 85.2% | 38.9% | 33.3% | 32.1% |

| | | | | | | |
|---|---|---|---|---|---|---|
| | non-sharps waste container | 100.0% | 96.3% | 94.4% | 48.1% | 67.9% |
| | sharps container | 100.0% | 100.0% | 88.9% | 94.4% | 96.4% |
| | environmental disinfectant | 90.9% | 100.0% | 72.2% | 94.4% | 96.4% |
| Medicines & commodities | Injectable uterotonic | 90.9% | 70.4% | 77.8% | 75.9% | 73.8% |
| | injectable beta/dexamethasone, | 100.0% | 100.0% | 33.3% | 14.8% | 22.6% |
| | IV solution and perfusion kit, | 81.8% | 70.4% | 83.3% | 55.6% | 67.9% |
| | misoprostol, | 45.5% | 85.2% | 66.7% | 9.3% | 28.6% |
| | oxygen and administration equipment, | 54.5% | 70.4% | 55.6% | 1.9% | 9.5% |
| | antibiotic eye ointment (tetracycline), | 63.6% | 81.5% | 88.9% | 59.3% | 66.7% |
| | injectable magnesium sulphate, | 100.0% | 88.9% | 72.2% | 24.1% | 39.3% |
| | injectable antibiotic for maternal/neonatal sepsis, | 81.8% | 100.0% | 100.0% | 94.4% | 89.3% |
| | skin disinfectant for cord care (chlorhexidine gel)* | | | 83.3% | 40.7% | 52.4% |
| | injectable Vitamin K, | 18.2% | 77.8% | 66.7% | 74.1% | 67.9% |
| | injectable calcium gluconate, | 36.4% | 81.5% | 61.1% | 22.2% | 23.8% |
| | Hypertensive (hydralazine, nifedipine, or methyldopa), | 36.4% | 85.2% | 77.8% | 20.4% | 33.3% |
| | injectable metronidazole | 18.2% | 92.6% | 72.2% | 79.6% | 73.8% |
| | injectable diazepam | 72.7% | 92.6% | NA | 81.5% | 78.6% |
| Human resources and guidelines | Staff trained in newborn resuscitation | 54.5% | 59.3% | 100.0% | 14.8% | 27.4% |
| | staff trained in essential childbirth care | 81.8% | 63.0% | 100.0% | 27.8% | 36.9% |
| | staff trained in essential newborn care | 90.9% | 66.7% | 94.4% | 37.0% | 52.4% |
| | guidelines for essential childbirth care | 36.4% | 40.7% | 16.7% | 37.0% | 35.7% |
| | checklist/job aids for essential childbirth care | 72.7% | 37.0% | 55.6% | 9.3% | 8.3% |
| | guidelines for essential newborn care | 90.9% | 44.4% | 22.2% | 27.8% | 39.3% |
| | skilled attendant present on call (onsite or in near proximity) 24 hours | 100.0% | 88.9% | 88.9% | 74.1% | 85.7% |
| Basic amenities | improved water source | 100.0% | 100.0% | 100.0% | 96.3% | 97.6% |
| | improved sanitation facility | 90.9% | 100.0% | 100.0% | 88.9% | 94.0% |
| | electricity available at all times when facility was open in last 7 days and/or back-up energy | 72.7% | 96.3% | 66.7% | 64.8% | 61.9% |
| | emergency transport (available on day of survey) | 54.5% | 66.7% | 44.4% | 33.3% | 34.5% |

| | | | | | | |
|---|---|---|---|---|---|---|
| Performance of BeMONC signal functions in last 12 months | Parenteral administration of antibiotics (IV or IM) for mothers | 90.9% | 92.6% | 100.0% | 94.4% | 97.6% |
| | Parenteral administration of anticonvulsants (magnesium sulfate) for management of pre-eclampsia and eclampsia (IV or IM) | 90.9% | 92.6% | 66.7% | 38.9% | 48.8% |
| | Parenteral administration of oxytocic for treatment of postpartum haemorrhage (IV or IM) | 90.9% | 96.3% | 94.4% | 94.4% | 96.4% |
| | Removal of retained products of conception using D&C or manual vacuum aspiration | 45.5% | 77.8% | 55.6% | 72.2% | 63.1% |
| | Assisted vaginal delivery using manual vacuum extraction (MVE) or forceps | 36.4% | 63.0% | 33.3% | 63.0% | 60.7% |
| | Manual removal of placenta | 90.9% | 96.3% | 88.9% | 94.4% | 91.7% |
| | Neonatal resuscitation with bag and mask | 90.9% | 100.0% | 77.8% | 46.3% | 45.2% |

Supplementary table 7. Facility readiness scores by facility type and managing authority.

| | | LUSAKA | | NAIROBI | OUAGADOUGOU | |
|---|---|---|---|---|---|---|
| | | Serving informal settlements (n=11)<br><br>%<br>SD | Not serving informal settlements (n=27)<br>%<br>SD | Serving informal settlements (n=18)<br><br>%<br>SD | Serving informal settlements (n=54)<br><br>%<br>SD | Not serving informal settlements (n=84)<br>%<br>SD |
| Labor & Delivery Care | Public Health Center | 66.1%<br>SD=6.8<br>*(n=7)* | *N/A* | 75.0%<br>SD=9.8<br>*(n=4)* | 57.8%<br>SD=11.9<br>*(n=18)* | 55.1%<br>SD=11.3<br>*(n=28)* |
| | Private Health Center | *N/A* | 86.5%<br>SD=9.1<br>*(n=19)* | 62.6%<br>SD=21.0<br>*(n=11)* | 54.2%<br>SD=12.4<br>*(n=24)* | 57.4%<br>SD=10.8<br>*(n=18)* |
| | Public Hospital | 86.6%<br>SD=8.4<br>*(n=4)* | 77.8%<br>SD=9.6<br>*(n=3)* | 87.7%<br>SD=3.8<br>*(n=3)* | 66.4%<br>SD=6.4<br>*(n=2)* | 64.0%<br>SD=13.0<br>*(n=10)* |
| | Private Hospital | *N/A* | 87.0%<br>SD=9.0<br>*(n=5)* | *N/A* | 54.5%<br>SD=11.5<br>*(n=10)* | 66.0%<br>SD=12.9<br>*(n=28)* |
| | *One-way ANOVA p- value** | ***p=0.002*** | *p=0.305* | *p=0.109* | *p=0.466* | ***p=0.005*** |
| Small & Sick Newborn Care | Public Health Center | 52.5%<br>SD=11.1<br>(n=7) | *N/A* | 64.2%<br>SD=10.5<br>*(n=4)* | 39.1%<br>SD=7.3<br>*(n=18)* | 37.5%<br>SD=10.2<br>*(n=28)* |
| | Private Health Center | *N/A* | 71.6%<br>SD=11.0<br>*(n=19)* | 56.0%<br>SD=20.0<br>*(n=11)* | 34.5%<br>SD=8.6<br>*(n=24)* | 35.1%<br>SD=9.2<br>*(n=18)* |
| | Public Hospital | 75.3%<br>SD=4.0 | 62.2%<br>SD=9.0 | 76.7%<br>SD=5.5 | 49.3%<br>SD=8.7 | 44.6%<br>SD=12.2 |

| | | | | | | |
|---|---|---|---|---|---|---|
| | | (n=4) | *(n=3)* | *(n=3)* | *(n=2)* | *(n=10)* |
| | Private Hospital | *N/A* | 72.1%<br>SD=14.2<br>*(n=5)* | *N/A* | 38.1%<br>SD=12.0<br>*(n=10)* | 44.9%<br>SD=14.6<br>*(n=28)* |
| | *One-way ANOVA p- value** | ***p=0.004*** | *p=0.411* | *p=0.202* | *p=0.095* | ***p=0.019*** |

**one-way analysis of variance (ANOVA) p<0.05 suggests that at least one of the mean scores is statistically significantly different, across facility types and managing authority.*

Supplementary figure 3. Labor and delivery readiness scores (%) by facility type and managing authority

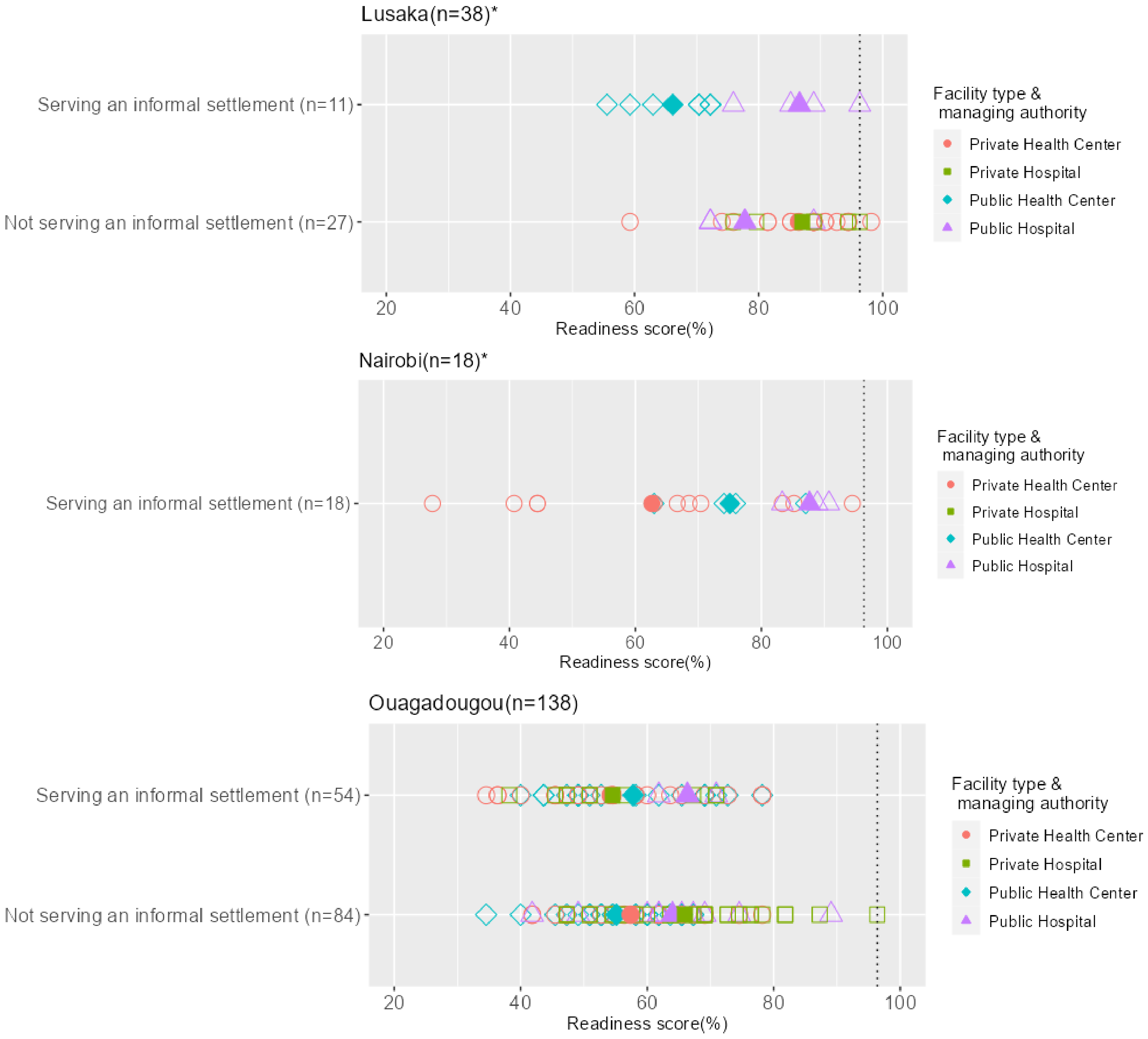

*Notes:*

**in Lusaka there are no private facilities serving informal settlements, and there are no public health centers in areas not serving informal settlements; in Nairobi there are no private hospitals serving informal settlements.*

*Filled symbols represent the average readiness score for a particular facility type/managing authority; hollow symbols indicate the readiness score for each facility.*

*Dotted line represents the maximum readiness score for lower-level facilities, based on exclusion of infant incubator and resuscitation table, and was calculated as 96.3% in Nairobi and Lusaka, and 96.4% in Ouagadougou.*

Supplementary figure 4. Distribution of small and sick newborn readiness scores comparing facilities serving and not serving informal settlements, by city

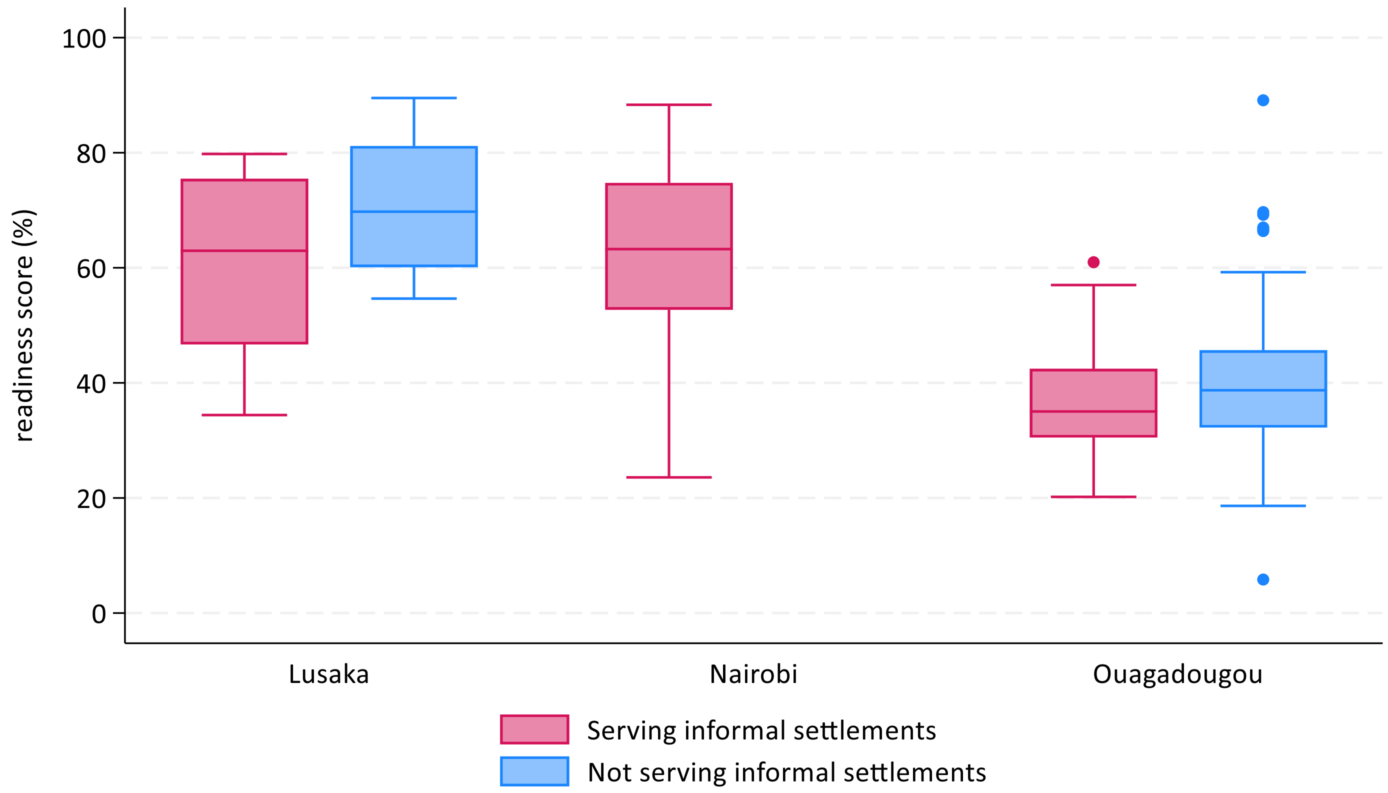

Supplementary figure 5 A-C: Radar plots of small & sick newborn care item availability by city.

A. Lusaka: facilities serving informal settlements (n=11)

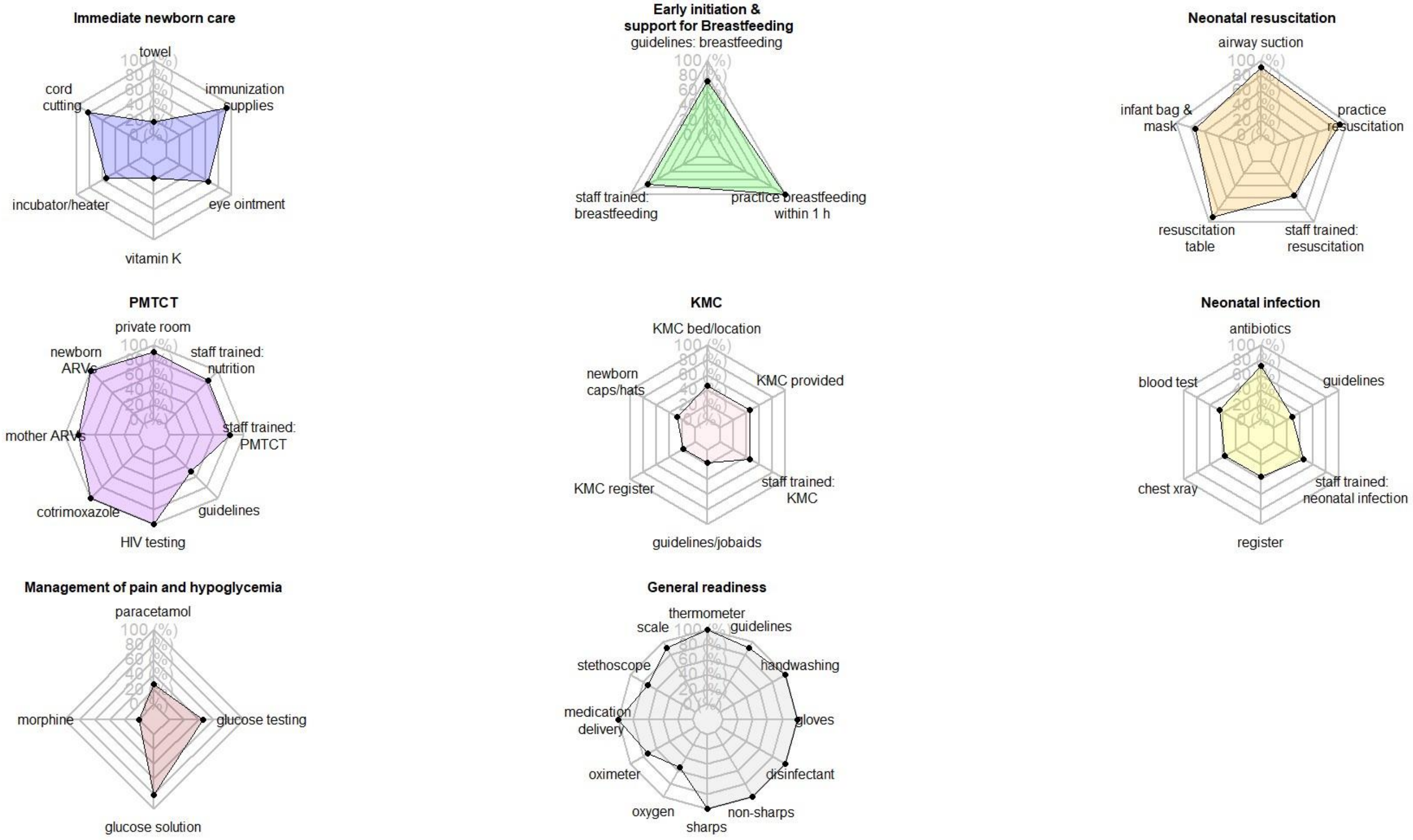

B. Nairobi: facilities serving study area (n=18)

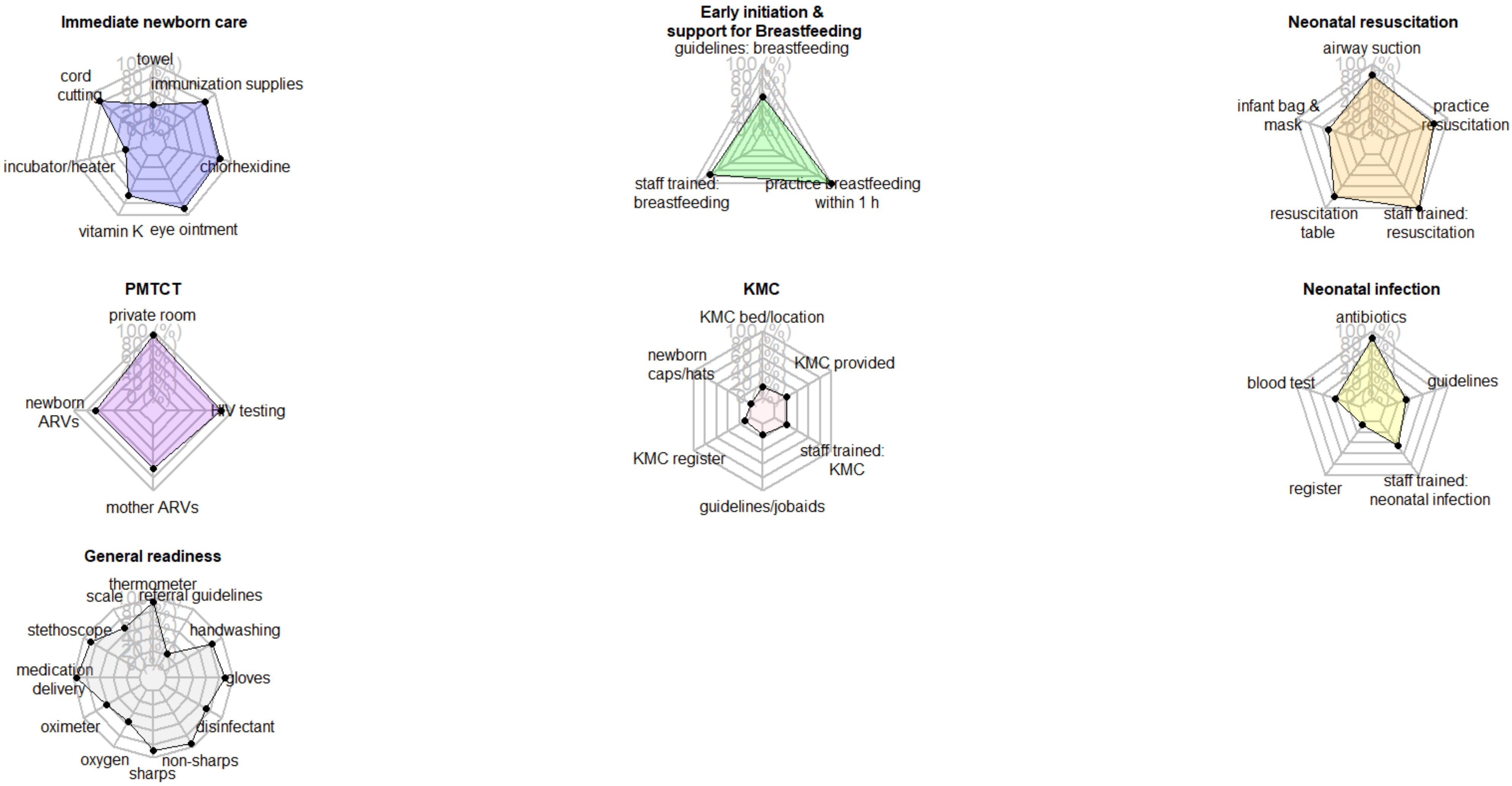

C. Ouagadougou: facilities serving informal settlements (n=54)

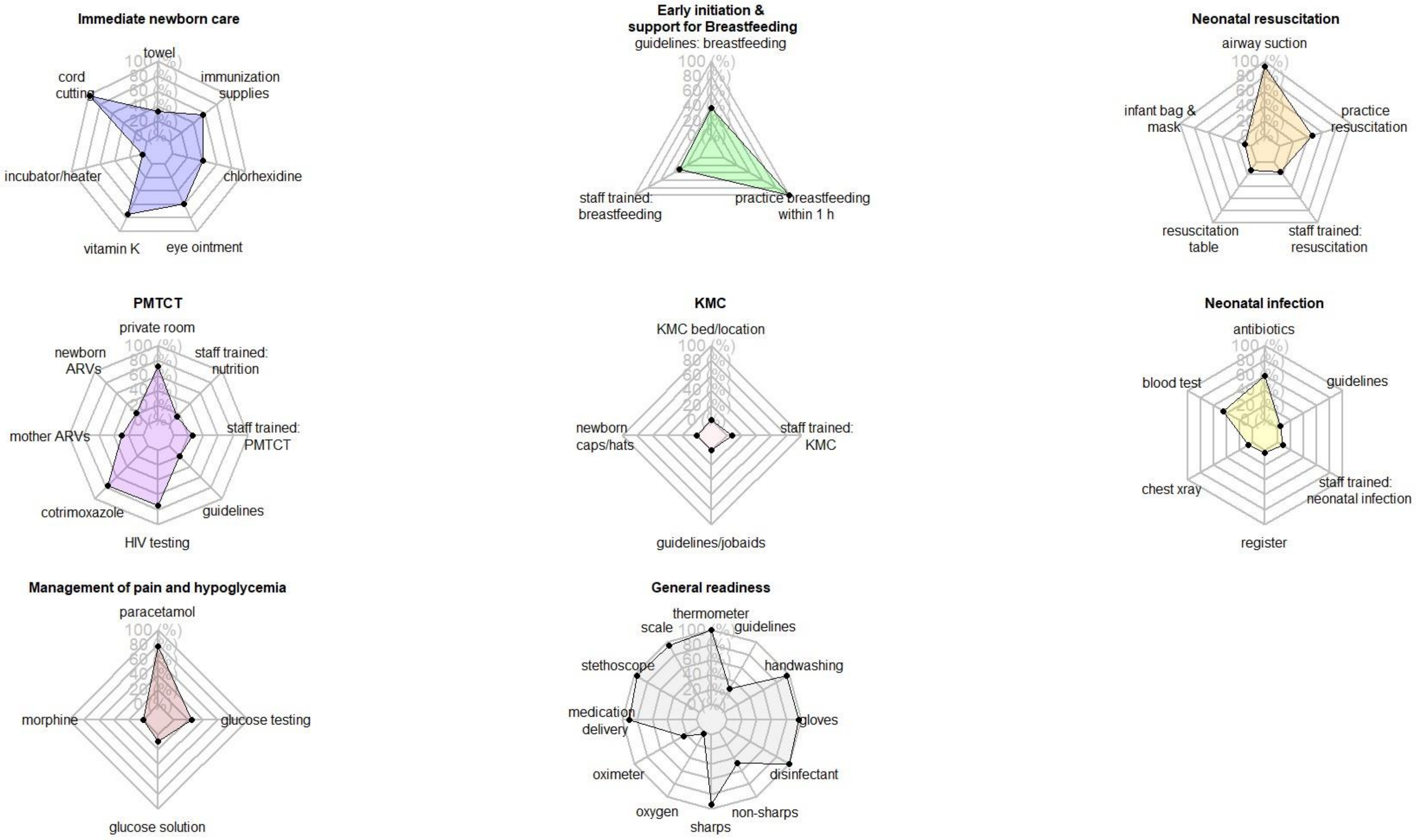

Supplementary table 8. Item availability for small & sick newborn care readiness by city and facilities serving/not serving informal settlements.

| | | LUSAKA | | NAIROBI | OUAGADOUGOU | |
|---|---|---|---|---|---|---|
| Domain | Item | Serving informal settlements (n=11) | Not serving informal settlements (n=27) | Serving informal settlements (n=18) | Serving informal settlements (n=54) | Not serving informal settlements (n=84) |
| Immediate newborn care and routine care | Clean towel for drying newborn, | 18.2% | 85.2% | 38.9% | 33.3% | 32.1% |
| | cord cutting supplies | 81.8% | 100.0% | 83.3% | 98.1% | 92.9% |
| | radiant heater/warmth source or infant incubator | 54.5% | 81.5% | 22.2% | 1.9% | 11.9% |
| | Vitamin K (inj) | 18.2% | 77.8% | 66.7% | 74.1% | 67.9% |
| | antibiotic eye ointment, | 63.6% | 81.5% | 88.9% | 59.3% | 66.7% |
| | chlorhexidine solution | | | 83.3% | 40.7% | 52.4% |
| | immunization supplies | 92.4% | 67.3% | 80.6% | 56.8% | 60.9% |
| Early initiation and support for breastfeeding | Guidelines for breastfeeding and promotion of breastfeeding, | 72.7% | 40.7% | 50.0% | 37.0% | 45.2% |
| | Staff trained in breastfeeding and promotion of breastfeeding, | 72.7% | 40.7% | 72.2% | 29.6% | 40.5% |
| | routine practice of breastfeeding initiation within 1 hour of birth | 100.0% | 100.0% | 100.0% | 100.0% | 98.8% |
| Neonatal resuscitation | Airway suction apparatus, | 90.9% | 96.3% | 83.3% | 92.6% | 92.9% |
| | infant resuscitation bag & mask, | 72.7% | 96.3% | 50.0% | 9.3% | 11.9% |
| | resuscitation table with heat source, | 90.9% | 100.0% | 77.8% | 13.0% | 25.0% |
| | Staff trained in neonatal resuscitation using bag and mask, | 54.5% | 59.3% | 100.0% | 14.8% | 27.4% |
| | Facility provided neonatal resuscitation in past three months | 90.9% | 100.0% | 77.8% | 46.3% | 45.2% |
| | PMTCT room is private room with auditory and visual privacy | 90.9% | 88.9% | 94.4% | 72.2% | 73.8% |

| | | | | | | |
|---|---|---|---|---|---|---|
| Prevention of mother-to-child transmission of HIV (PMTCT) | Antiretrovirals for newborns | 100.0% | 81.5% | 66.7% | 22.2% | 31.0% |
| | Antiretrovirals for mothers | 81.8% | 37.0% | 66.7% | 29.6% | 33.3% |
| | Cotrimoxazole | 100.0% | 96.3% | N/A | 75.9% | 71.4% |
| | HIV diagnostic capacity | 100.0% | 92.6% | 83.3% | 74.1% | 79.8% |
| | PMTCT and infant and young child feeding (IYCF) guidelines | 50.0% | 42.6% | N/A | 19.4% | 27.4% |
| | Staff trained PMTCT in last 2 years | 81.8% | 55.6% | N/A | 25.9% | 17.9% |
| | Staff trained in newborn nutrition counseling of mother with HIV or IYCF in last years. | 81.8% | 70.4% | N/A | 14.8% | 13.1% |
| Kangaroo Mother Care (KMC) | bed/location for caregiver to stay overnight to provide KMC | 45.5% | 37.0% | 16.7% | 0.0% | 2.4% |
| | caps/hats for newborn | 27.3% | 33.3% | 0.0% | 0.0% | 3.6% |
| | register to record KMC services | 18.2% | 7.4% | 11.1% | N/A | N/A |
| | Guidelines, protocols or job aids for KMC, | 18.2% | 29.6% | 16.7% | 0.0% | 3.6% |
| | Staff trained in KMC in last 2 years, | 45.5% | 33.3% | 22.2% | 7.4% | 6.0% |
| | facility provided KMC in last 3 months. | 45.5% | 22.2% | 22.2% | N/A | N/A |
| Detection and management of neonatal infection | First line antibiotics for neonatal sepsis OR second line antibiotics for neonatal sepsis, | 72.7% | 100.0% | 88.9% | 59.3% | 69.0% |
| | full blood count diagnostic capacity, | 45.5% | 66.7% | 38.9% | 44.4% | 45.2% |
| | chest x-ray capacity | 36.4% | 63.0% | N/A | 5.6% | 2.4% |
| | register for neonatal sepsis | 36.4% | 14.8% | 5.6% | 3.7% | 9.5% |
| | staff trained in neonatal sepsis in last 2 years | 45.5% | 33.3% | 44.4% | 7.4% | 13.1% |
| | guidelines/checklists for neonatal sepsis | 27.3% | 33.3% | 33.3% | 3.7% | 9.5% |
| Comfort and pain management | Paracetamol syrup | 27.3% | 88.9% | N/A | 77.8% | 65.5% |
| | morphine (syrup or injectable) | 0.0% | 44.4% | N/A | 0.0% | 6.0% |
| Detection and management of hypoglycemia | Glucose injectable solution, | 81.8% | 100.0% | N/A | 9.3% | 7.1% |
| | Blood glucose testing capacity. | 45.5% | 88.9% | N/A | 24.1% | 33.3% |
| General facility readiness | Thermometer, | 100.0% | 100.0% | 94.4% | 100.0% | 100.0% |
| | infant scale, | 90.9% | 100.0% | 66.7% | 94.4% | 97.6% |
| | stethoscope, | 72.7% | 96.3% | 88.9% | 96.3% | 97.6% |
| | medication delivery mechanism, | 100.0% | 100.0% | 94.4% | 90.7% | 94.0% |
| | pulse oximeter, | 72.7% | 100.0% | 61.1% | 24.1% | 39.3% |

| | | | | | | |
|---|---|---|---|---|---|---|
| | oxygen and administration equipment, | 54.5% | 70.4% | 55.6% | 1.9% | 9.5% |
| | non-sharps waste container, | 100.0% | 96.3% | 94.4% | 48.1% | 67.9% |
| | Sharps container, | 100.0% | 100.0% | 88.9% | 94.4% | 96.4% |
| | environmental disinfectant, | 100.0% | 100.0% | 72.2% | 100.0% | 98.8% |
| | sterile gloves, | 100.0% | 100.0% | 88.9% | 96.3% | 98.8% |
| | handwashing materials, | 100.0% | 100.0% | 83.3% | 96.3% | 94.0% |
| | guidelines for referral of small or sick newborn | 90.9% | 44.4% | 22.2% | 27.8% | 39.3% |

Supplementary figure 6. Small and sick newborn care readiness scores (%) by facility type and managing authority

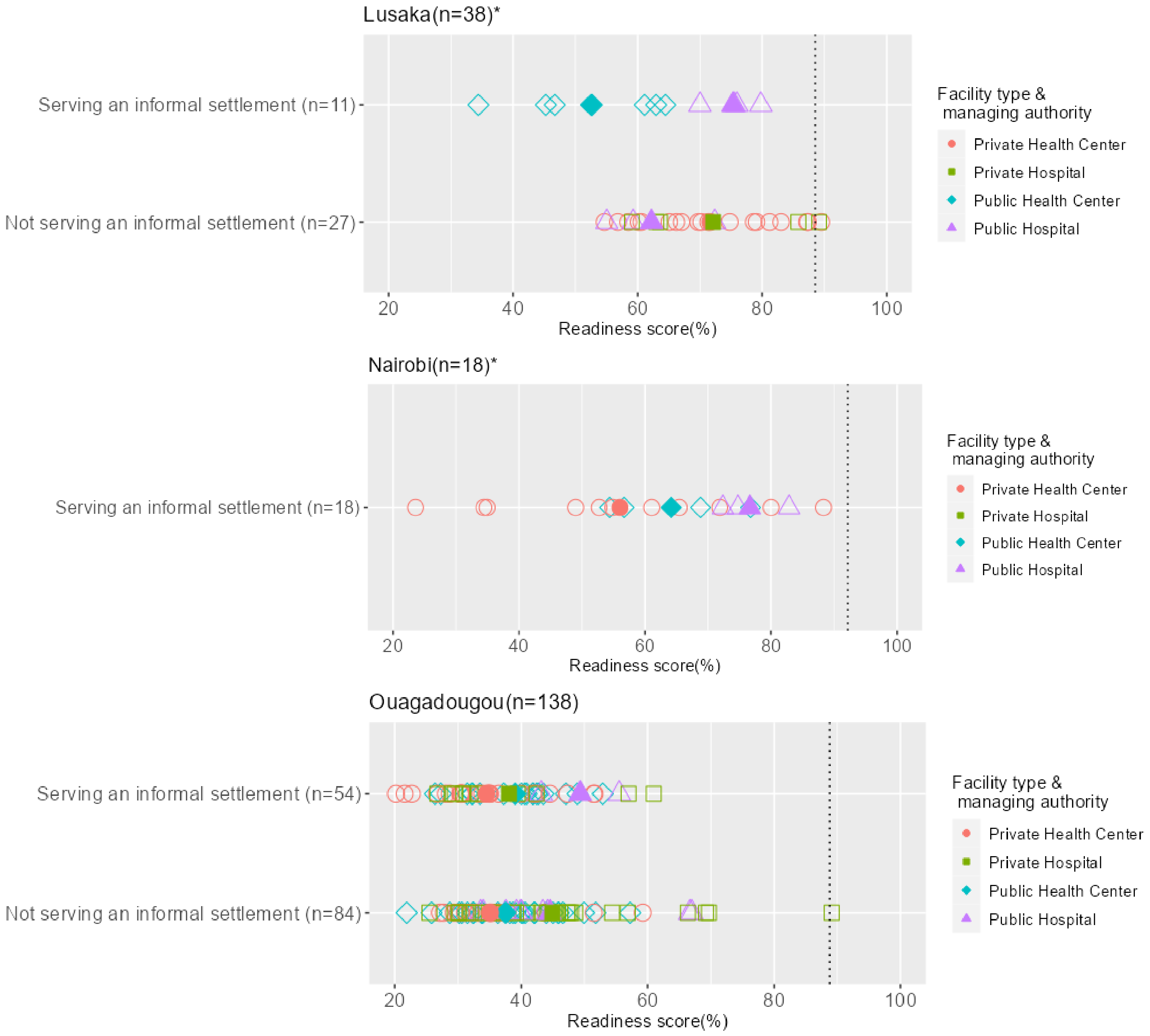

*Notes:*

**in Lusaka there were no private facilities serving informal settlements, and there are no public health centers in areas not serving informal settlements; in Nairobi there were no private hospitals serving informal settlements.*

*Filled symbols represent the average readiness score for a particular facility type/managing authority; hollow symbols indicate the readiness score for each facility.*

*Dotted line represents the maximum score for lower-level facilities, based on exclusion of infant incubator, resuscitation table, x-ray machine, and morphine, and was calculated at 88.5% in Lusaka, 92.2% in Nairobi and 88.8% in Ouagadougou.*

Supplementary table 9. Characteristics of women included in exact match linking analysis in Nairobi, Lusaka and Ouagadougou.

| | Lusaka (n=424) | | | Nairobi (n=411) | | | Ouagadougou (n=401) | | |
|---|---|---|---|---|---|---|---|---|---|
| Women's characteristics | % | SD | n | % | SD | n | % | SD | n |
| *Age group* | | | 424 | | | 411 | | | 401 |
| 15-19 yrs | 11.1% | 31.4 | | 6.1% | 23.9 | | 6.7% | 25.1 | |
| 20-34 yrs | 76.9% | 42.2 | | 82.7% | 37.8 | | 80.0% | 40.0 | |
| 35-49 yrs | 12.0% | 32.6 | | 11.2% | 31.6 | | 13.2% | 33.9 | |
| *Education category* | | | 424 | | | 411 | | | 401 |
| None | 2.8% | 16.6 | | 6.6% | 24.8 | | 42.1% | 49.4 | |
| Primary | 25.5% | 43.6 | | 24.6% | 43.1 | | 25.4% | 43.6 | |
| Secondary + | 71.7% | 45.1 | | 68.9% | 46.4 | | 32.4% | 46.9 | |
| *Marital status* | | | | | | | | | |
| In union | 79.0% | 40.7 | | 87.1% | 33.5 | | 98.5% | 12.1 | |
| Not in union | 21.0% | 40.7 | | 12.9% | 33.5 | | 1.5% | 12.1 | |
| *Employment status* | | | 424 | | | 411 | | | 401 |
| Not employed/no income | 57.3% | 49.5 | | 63.0% | 48.3 | | 48.6% | 50.0 | |
| Employed (public, private, self) | 33.7% | 47.3 | | 23.1% | 42.2 | | 17.2% | 37.8 | |
| Informal/casual labor | 9.0% | 28.6 | | 13.9% | 34.6 | | 34.2% | 47.5 | |
| *Parity* | | | 423 | | | 411 | | | 401 |
| 1 | 27.7% | 44.8 | | 28.7% | 45.3 | | 19.7% | 39.8 | |
| 2-3 | 49.9% | 50.1 | | 59.6% | 49.1 | | 52.6% | 50.0 | |
| 4+ | 22.5% | 41.8 | | 11.7% | 32.2 | | 27.7% | 44.8 | |
| *Facility type & managing authority* | | | 424 | | | 411 | | | 401 |
| Public Health Center | 45.8% | 49.9 | | 23.8% | 42.7 | | 52.6% | 50.0 | |
| Private Health Center | | | | 23.4% | 42.4 | | 17.0% | 37.6 | |
| Public Hospital | 54.2% | 49.9 | | 52.8% | 50.0 | | 13.2% | 33.9 | |
| Private Hospital | | | | 0.0% | | | 17.2% | 37.8 | |
| *Number of ANC contacts* | | | 424 | | | 410 | | | 380 |

| | | | | | | | | | |
|---|---|---|---|---|---|---|---|---|---|
| 0 | 1.4% | 11.8 | | 1.0% | 9.8 | | 2.6% | 16.0 | |
| 1-3 | 30.0% | 45.9 | | 32.7% | 47.0 | | 28.7% | 45.3 | |
| 4+ | 68.6% | 46.5 | | 66.3% | 47.3 | | 68.7% | 46.4 | |
| *Provider assisting at birth (highest level reported)* | | | 424 | | | 411 | | | 401 |
| Physician/specialist | 10.1% | 30.2% | | 43.3% | 49.6 | | 2.5% | 15.6 | |
| Midwife, TBA, nurse | 86.1% | 34.7% | | 54.7% | 49.8 | | 55.1% | 49.8 | |
| Other/unskilled | 0.9% | 9.7% | | | | | 0.5% | 7.1 | |
| Don't Know | 2.8% | 16.6% | | 1.9% | 13.8 | | 41.9% | 49.4 | |
| *Private bed for labor/delivery* | | | 424 | | | 411 | | | 401 |
| No | 14.6% | 35.4% | | 11.7% | 32.2 | | 10.0% | 30.0 | |
| Yes | 85.4% | 35.4% | | 88.3% | 32.2 | | 87.3% | 33.4 | |
| Don't know | | | | | | | 2.7% | 16.4 | |
| *Accompanied by partner/family during labor/delivery* | | | 424 | | | 411 | | | 401 |
| No | 98.3% | 12.8% | | 95.9% | 19.9 | | 57.9% | 49.4 | |
| Yes | 1.7% | 12.8% | | 4.1% | 19.9 | | 39.7% | 49.0 | |
| Don't know | | | | | | | 2.5% | 15.6 | |
| Length of stay in facility | | | 424 | | | 411 | | | 401 |
| <24h | 73.3% | 44.3% | | 16.1% | 36.8 | | 65.8% | 47.5% | |
| ≥24h | 26.7% | 44.3% | | 83.9% | 36.8 | | 34.2% | 47.5% | |
| *Maternal PNC check before discharge* | | | 419 | | | 410 | | | 398 |
| No | 5.5% | 22.8% | | 4.4% | 20.5 | | 10.1% | 30.1 | |
| Yes | 94.5% | 22.8% | | 95.6% | 20.5 | | 89.9% | 30.1 | |
| *Newborn PNC check before discharge* | | | 419 | | | 410 | | | 399 |
| No | 8.1% | 27.3% | | 5.9% | 23.5 | | 12.8% | 33.4 | |
| Yes | 91.9% | 27.3% | | 94.1% | 23.5 | | 87.2% | 33.4 | |

Supplementary table 10. Labor and delivery readiness scores (%) in facilities included in exact match linking analysis in Nairobi, Lusaka and Ouagadougou.

| Domain | LUSAKA<br>Serving study area<br>(n=10) | NAIROBI<br>Serving study area<br>(n=16) | OUAGADOUGOU<br>Serving study area<br>(n=54) |
|---|---|---|---|
| Overall labor & delivery readiness *(simple additive score)* | 73.7% | 72.7% | 59.8% |
| Equipment | 79.1% | 67.9% | 64.1% |
| Medicines & commodities | 60.8% | 76.9% | 50.3% |
| Basic amenities | 80.0% | 79.7% | 72.2% |
| Human resources & guidelines | 74.3% | 71.4% | 44.2% |
| Performance of BeMONC signal function in last 12 months | 75.7% | 77.7% | 73.3% |
| *% facilities with score >=95% (ceiling)* | *10.0%* | *0.0%* | *0.0%* |
| *% facilities with score <=5% (floor)* | *0.0%* | *0.0%* | *0.0%* |

Supplementary table 11. Associations between facilities' labor & delivery care readiness scores (%) and women's PCMC domain scores in study areas by city.

| | Crude model | | | Adjusted Model | | |
|---|---|---|---|---|---|---|
| | Coefficient [95%CI] | P value | n | Coefficient [95%CI] | P value | n |
| **DIGNITY AND RESPECT (18 points)** | | | | | | |
| Lusaka[+] | -0.03 [-0.08, 0.03] | 0.299 | 424 | -0.02 [-0.07, 0.03] | 0.370 | 410 |
| Nairobi[++] | 0.01 [-0.03, 0.05] | 0.684 | 411 | 0.01 [-0.02, 0.04] | 0.376 | 406 |
| Ouagadougou[+++] | 0.02 [-0.02, 0.06] | 0.265 | 401 | 0.03 [-0.01, 0.07] | 0.152 | 373 |
| **COMMUNICATION & AUTONOMY (27 points)** | | | | | | |
| Lusaka+ | -0.003 [-0.13, 0.12] | 0.965 | 424 | -0.07 [-0.29, 0.15] | 0.536 | 410 |
| Nairobi[++] | -0.06 [-0.14, 0.15] | 0.115 | 411 | **-0.06 [-0.12, -0.01]** | **0.027** | 407 |
| Ouagadougou[+++] | -0.01 [-0.07, 0.05] | 0.732 | 401 | 0.01 [-0.05, 0.07] | 0.723 | 371 |
| **SUPPORTIVE CARE (45 points)** | | | | | | |
| Lusaka[+] | -0.11 [-0.23, 0.01] | 0.081 | 424 | **-0.22 [-0.37, -0.07]** | **0.004** | 408 |
| Nairobi[++] | -0.01 [-0.13, 0.11] | 0.869 | 411 | **0.08 [0.01, 0.15]** | **0.030** | 406 |
| Ouagadougou[+++] | 0.04 [-0.05, 0.14] | 0.381 | 401 | 0.04 [-0.05, 0.13] | 0.407 | 394 |

*All adjusted models included variables significant at p<0.2 in bivariate association with PCMC domain scores:*

+ Lusaka:

*Dignity and respect model adjusted by maternal PNC check received before discharge, PNC counseling on danger signs received before discharge, PNC, blood pressure checked before discharge, newborn PNC check received before discharge, appointment received for next PNC check before discharge; Communication and autonomy model adjusted by: facility type and managing authority, women's education, women's employment status, women's age, history of pregnancy complications, type of provider assisting during delivery, maternal PNC check received before discharge, PNC counseling on danger signs received before discharge, PNC counseling on family planning received before discharge, blood pressure checked before discharge, newborn PNC check received before discharge; Supportive care model adjusted by: facility type and managing authority, parity, type of provider assisting during delivery, maternal PNC check received before discharge, PNC counseling on danger signs received before discharge, PNC counseling on family planning received before discharge, blood pressure checked before discharge, newborn PNC check received before discharge, appointment received for next PNC check before discharge.*

++ Nairobi:

*Dignity and respect model adjusted by delivery facility type and managing authority, women's employment, women's age, type of provider assisting during delivery, maternal PNC check received before discharge, PNC counseling on danger signs received before discharge, blood pressure checked before discharge, newborn PNC check received before discharge, appointment received for next PNC check before discharge; Communication and autonomy model adjusted by: facility type and managing authority, women's employment status, women's marital status, place of ANC, type of provider assisting during delivery, length of facility stay, PNC counseling on danger signs received before discharge, PNC counseling on family planning received before discharge, blood pressure checked before discharge, newborn PNC check received before discharge; Supportive care model adjusted by: facility type and managing authority, women's education, women's age, marital status, type of provider assisting during delivery, maternal PNC check received before discharge, PNC counseling on danger signs received before discharge, PNC counseling on family planning received before discharge, blood pressure checked before discharge, newborn PNC check received before discharge, appointment received for next PNC check before discharge.*

+++ Ouagadougou:

*Dignity and respect model adjusted by delivery facility type and managing authority, women's marital status, parity, ANC4+, type of provider assisting during delivery, maternal PNC check received before discharge, PNC counseling on danger signs received before discharge, PNC counseling on family planning received before discharge, blood pressure checked before discharge, newborn PNC check received before discharge, appointment received for next PNC check before discharge; Communication and autonomy model adjusted by: facility type and managing authority, women's employment status, history of pregnancy complications, ANC4+, type of provider assisting during delivery, maternal PNC check received before discharge, PNC counseling on danger signs received before discharge, PNC counseling on family planning received before discharge, blood pressure checked before discharge, newborn PNC check received before discharge, appointment received for next PNC check before discharge; Supportive care model adjusted by: facility type and managing authority, history of miscarriage/stillbirth, type of provider assisting during delivery, maternal PNC check received before discharge, length of stay in facility, PNC counseling on danger signs received before discharge, PNC counseling on family planning received before discharge, blood pressure checked before discharge, newborn PNC check received before discharge.*
